# Outburst of serotype 4 IPD after COVID-19 is driven by ST15063/GPSC162 lineage associated with high-risk behaviors and greater virulence linked to influenza H3N2 virus coinfection and cigarette smoke

**DOI:** 10.64898/2026.02.27.26346872

**Authors:** Covadonga Pérez-García, Joaquin Llorente, Maria Elena Aguirre Alastuey, Mirella Llamosí, Ruth Gil, Gabriel Laghlali, Farah El-Ayache, Vivian Yan, Michael Schotsaert, Jorge del Diego, Jose Miguel Cisneros, Adolfo García-Sastre, Mirian Domenech, Julio Sempere, Jose Yuste

## Abstract

The emergence of vaccine covered serotypes causing invasive pneumococcal disease (IPD) is a serious concern worldwide. We investigated the unexpected rise of serotype 4 causing IPD primarily in non-vaccinated young adults after the COVID-19 pandemic that further spread to adults ≥ 65 years in recent years. For this purpose, we conducted a retrospective study of serotype 4 IPD cases (n=827) reported in Spain between 2009 and 2024. Whole-genome sequencing was performed to assess clonal lineages and phylogenetic relationships. Clinical and epidemiological data were compared between serotype 4 and all other serotypes causing IPD. Epidemiological and genomic analysis confirmed that the rise started as an abrupt cluster of IPD cases in Seville (Andalusia) in the year 2022 due to the ST15063 within GPSC12 lineage. This outbreak initially caused pneumonia episodes that required hospitalization in young individuals associated with high rates of tobacco smoking, alcohol, and inhaled drugs such as cannabis and cocaine, followed by a general distribution pattern throughout the country in the following years, affecting the elderly population. Experimental studies to evaluate potential underlying mechanisms confirmed that ST15063 serotype 4 strains displayed enhanced infection rates of human lung cells that significantly increased in the presence of cigarette smoke exposure and by influenza H3N2 virus coinfection, but not with H1N1. These findings highlight the need for targeted vaccination strategies not only against pneumococcus but also against respiratory viruses such as influenza, RSV and COVID-19 and demonstrate the importance of molecular surveillance to establish effective interventions in high-risk populations.

## Introduction

Invasive pneumococcal disease (IPD) remains a significant public health concern worldwide due to its high morbidity and mortality rates, particularly among young children, older adults, and immunocompromised individuals. In Spain, the use of pneumococcal conjugate vaccines (PCVs) in children started in 2001 with PCV7, replaced in 2010 with PCV13 in the private market, and finally introduced in the national pediatric calendar in 2016 with high coverage rates >95%. The use of these vaccines clearly reduced the burden of disease including antibiotic resistance with a clear impact in adult IPD due to their herd-immune effects [1–3]. However, serotype replacement by non-PCV13 serotypes has been increasingly reported in recent years, especially serotype 8 [2,4,5], and since COVID-19 pandemic, cases by PCV13 serotypes such as serotype 3 have also spiked in both adult and pediatric populations [4].

While serotype 4 has historically been uncommon in adult IPD, recent surveillance data indicate a notable rise in IPD cases caused by this vaccine serotype, especially among Spanish adults aged 18–64 years from ranking ninth in 2019, to third cause of IPD by 2022–2023 [4]. This pattern has also been observed in other countries, for example, in the UK, IPD by serotype 4 was scarce in previous seasons, but cases surged during 2022-2023 in young adults [6]. In Canada, serotype 4 was also one of the main causes of the resurgence of IPD cases in adults after the COVID-19 pandemic [7]. However, other countries have only linked serotype 4 cases to specific high-risk groups, such as individuals experiencing homelessness or opioid use [8,9], or specific outbreaks in workers, such as in shipyards in Northern Europe [10].

Given the changing epidemiology of serotype 4 and its presence or not in higher-valency PCVs (PCV20 or PCV21 respectively), understanding its recent emergence is essential to update vaccine policy and IPD prevention strategies. This study analyzes serotype 4 IPD trends in Spain from 2009 to 2024, performing a WGS-analysis of almost 400 serotype 4 isolates to analyze main circulating genetic lineages. Moreover, we describe the initial location of the outburst of serotype 4 cases in young adults, with complete clinical data of patients suffering from IPD by this emerging serotype. In addition, experimental assays were performed to evaluate the impact of cigarette smoke exposure in bacterial adhesion to the lung, and the contribution of influenza virus co-infections to the rise of vaccine serotypes such as serotype 4.

## Methods

### Study design

We characterized IPD isolates affecting all age groups submitted by hospital microbiology laboratories to the Spanish Pneumococcal Reference Laboratory (SPRL). The SPRL reports every year to the European Center for Disease Control (ECDC) all IPD cases in Spain and to the Invasive Respiratory Infection Surveillance Network (IRIS) [11,12]. Serotyping was performed using Quellung reaction, dot blot assay using specific antisera, and/or PCR-capsular sequence typing [2,4]. For this study, we considered all serotype 4 isolates received at the laboratory covering the period 2009 to 2024 (n=827).

The epidemiological year considered in the manuscript is from January to December. IPD trends were analyzed for different age groups covering the pediatric population (<2 years, 2–4 years, 5–17 years) and adult population (18–64 years, ≥65 years). Isolates from 2009-2024 were divided in different vaccine periods (pre-PCV13, 2009; early PCV13, 2010-12; middle PCV13, 2013-2016; late PCV13, 2017-2019; COVID-19, 2020-21; reopening, 2022-23; new PCVs, 2024). Due to the specific location of serotype 4 cases in Spain, we also represent cases by geographical regions (Spanish Autonomous Communities or CCAA). Spain is divided into 17 autonomous communities (CCAA) or regions and 2 autonomous cities. These CCAA/regions are divided into different subregions known as provinces. Andalusia (AND) is a CCAA/region in southern Spain, and Seville (SEV) is a province/subregion of Andalusia (Supplemental Figure S1). For the whole genome sequencing (WGS) study, we sequenced 60-96% of isolates from years 2009 (pre-PCV13), 2015 (middle PCV13), 2018-19 (late PCV13), 2021 (COVID-19), 2022-23 (reopening), and 2024 (new PCVs).

### Whole genome sequencing and bioinformatic analysis

Chromosomal DNA was extracted using the Wizard Genomic DNA Purification Kit (Promega). Sequencing libraries were prepared with the Illumina DNA Prep (96) kit and IDT for Illumina Nextera DNA Unique Dual Indexes, followed by paired-end sequencing (2 × 150 bp) on a NovaSeq 6000 platform at the Genomics Unit, ISCIII. The resulting reads have been deposited in the European Nucleotide Archive (Supplemental Table S1).

WGS analysis was conducted using the Bactopia Nextflow pipeline [13]. The workflow is detailed in supplemental methods. All isolates were classified in sequence-type (ST) based upon the 7-locus multilocus sequence typing (MLST) scheme. CCs cluster pneumococci based upon the ST [14]. GPSCs define pneumococcal clusters based upon the analysis of core and accessory genes [15]. For serotype 4 phylogenetic analysis, reads were mapped to the ST801/CC801/GPSC162 reference genome described by Gladstone et al. (2022) (GCA_09128732) [10]. The reference was prepared in Geneious Prime (Biomatters) by removing contigs <500 bp, reordering the remaining contigs with Mauve alignment against the complete ATCC700669 genome, and concatenating them into a final sequence of 2,051,094 bp.

Geneious R9 (Biomatters) was also used to investigate mutations and gene acquisitions associated with AMR and to examine major virulence factors in serotype 4 isolates. Virulence determinants were identified using reference annotations from the TIGR4, D39, R6, OXC141, and ATCC700669 genomes. Prophages were classified into PPH families [16], and integrative and conjugative elements (ICEs) were detected using ICEberg 3.0 (https://tool2-mml.sjtu.edu.cn/ICEberg3/). Global phylogenetic contextualization and GPSC lineage assignments were obtained from PathogenWatch (https://pathogen.watch/).

### Clinical data from patients hospitalized with IPD by serotype 4 or other serotypes

Due to the specific expansion of serotype 4 causing IPD among young adults in the subregion of Seville, (Andalusia, Spain) since 2022, we evaluated clinical and demographic data of young adults (18–65 years) diagnosed with IPD by any serotype between January 2022 and December 2024 from the five main hospitals in Seville: Hospital Universitario Virgen del Rocío, Hospital Universitario Virgen de Macarena, Complejo Hospitalario Nuestra Señora de Valme, Hospital San Juan de Dios del Aljarafe, and Hospital de la Merced. All data were anonymized, and the study was approved by the Ethics Committee of the Hospital Universitario Virgen del Rocío (ENISE4, SICEIA-2025-002346).

Demographic variables included age, sex, postal code type (categorized as: home address with valid postal code, street homelessness, or penitentiary). Substance consumption included smoking status, alcohol, parenteral drug use, inhaled drug use and polydrug use. Comorbidities comprised chronic obstructive pulmonary disease (COPD), asthma, severe COVID-19, other chronic respiratory diseases, HIV infection, immunosuppression (of any cause), chronic corticosteroid use, arterial hypertension, diabetes mellitus, and dyslipidemia. Vaccination status was recorded for both COVID-19 and pneumococcal vaccines (categorized as no vaccination, PCV13, or PCV13+PPV23). Mortality during hospital admission (exitus) was also recorded.

### Effect of cigarette smoke and co-infection with influenza viruses in pneumococcal infection of human lung epithelial cells

Pneumococcal clinical isolates and influenza virus strains used for infection studies are listed in Supplemental Table S2, and a more detailed description of the adhesion assays is provided in the supplemental methods. Cigarette smoke extract (CSE) was prepared from research cigarettes (nicotine 0.73 mg, tar 9.4 mg, CO 12 mg) supplied by the University of Kentucky, as previously described [17], and stored at −80°C as 100% CSE [17].

Pneumococcal adhesion assays were performed using Detroit 562 (ATCC CCL-138) and A549 lung cells (CCL-185; ATCC) [17]. Cells were seeded at 2 × 10⁵ per well in 24-well plates and grown to ∼95% confluence. For adhesion assays, monolayers were washed with 1× PBS and infected with *S. pneumoniae* at the indicated multiplicity of infection (MOI) (25:1) for 1 h at 37°C with 5% CO₂. For co-infection studies, A549 cells in DMEM supplemented with 0.2% BSA, 1 mM MgCl₂, and 0.9 mM CaCl₂ were infected with influenza A/New Caledonia/20/1999 (H1N1) or A/Darwin/9/2021 (H3N2) at MOI 0.5 for 1 h. [18].

### Statistical Analysis

The corrected incidences were calculated as the number of IPD episodes per 100,000 population and year using population data from the Spanish National Statistical Institute and the Institute of Statistics of Andalusia as the denominator and considering the population capture of 80% [4]. We assumed the same epidemiological characteristics for the population suffering from IPD covered by hospitals and the general population. Comparison of different periods was analyzed by calculating the IRRs using Poisson regression models. Statistical analyses were performed using STATA v.14.

For the clinical data statistical analysis, the primary comparison was between patients infected by serotype 4 versus patients infected by all other serotypes. Categorical variables between groups were compared using Pearson’s Chi-squared test when all expected cell counts were high, and Fisher’s exact test was applied if Chi-square test assumptions were violated. All analysis were performed in R version 4.3.2.

All experiments including human lung epithelial cells were performed at least three times including independent replicates. Statistical analyses were conducted using two-tailed Student’s t-test or one-way ANOVA with Dunnett’s post hoc test, with significance defined as *P* < 0.05 (*), P < 0.01 (**), and *P* < 0.001 (***).

## Results

### Expansion of serotype 4 cases in young adults since the COVID-19 pandemic

We analyzed serotype 4 cases and incidence rates from 2009 to 2024 across different vaccine periods. PCV13 was initially approved for pediatric vaccination in Spain in 2010 as private use although reaching 67–82% pediatric coverage [4]. In 2016, it was included into the national pediatric schedule (2+1 regimen) and used in some regions for immunocompetent adults ≥65 years and also for high-risk adults, following a sequential PCV13+PPV23 schedule. Pediatric coverage remained high (95–97%) since 2016, even during the pandemic, while adult coverage was low (PCV13: 23–29%, PPV23: 18–26%). Throughout 2024, some regions changed to PCV15 or PCV20 for children and PCV20 for immunocompetent adults.

Serotype 4 cases in children were scarce even before PCV13 introduction with a total of 8 cases in all the pediatric population in 2009 (Figure 1A-B). IPD cases diminished after vaccine introduction, showing only two IPD cases by serotype 4 in the late PCV13 period (2017-2019) considering all group ages. Lack of serotype 4 cases in children was reported during COVID-19, although a moderate upsurge was found during 2022-2024 with 5 cases in children > 5 years old and only one case in the age group < 5 years old (Figure 1A-B).

**Figure 1.**
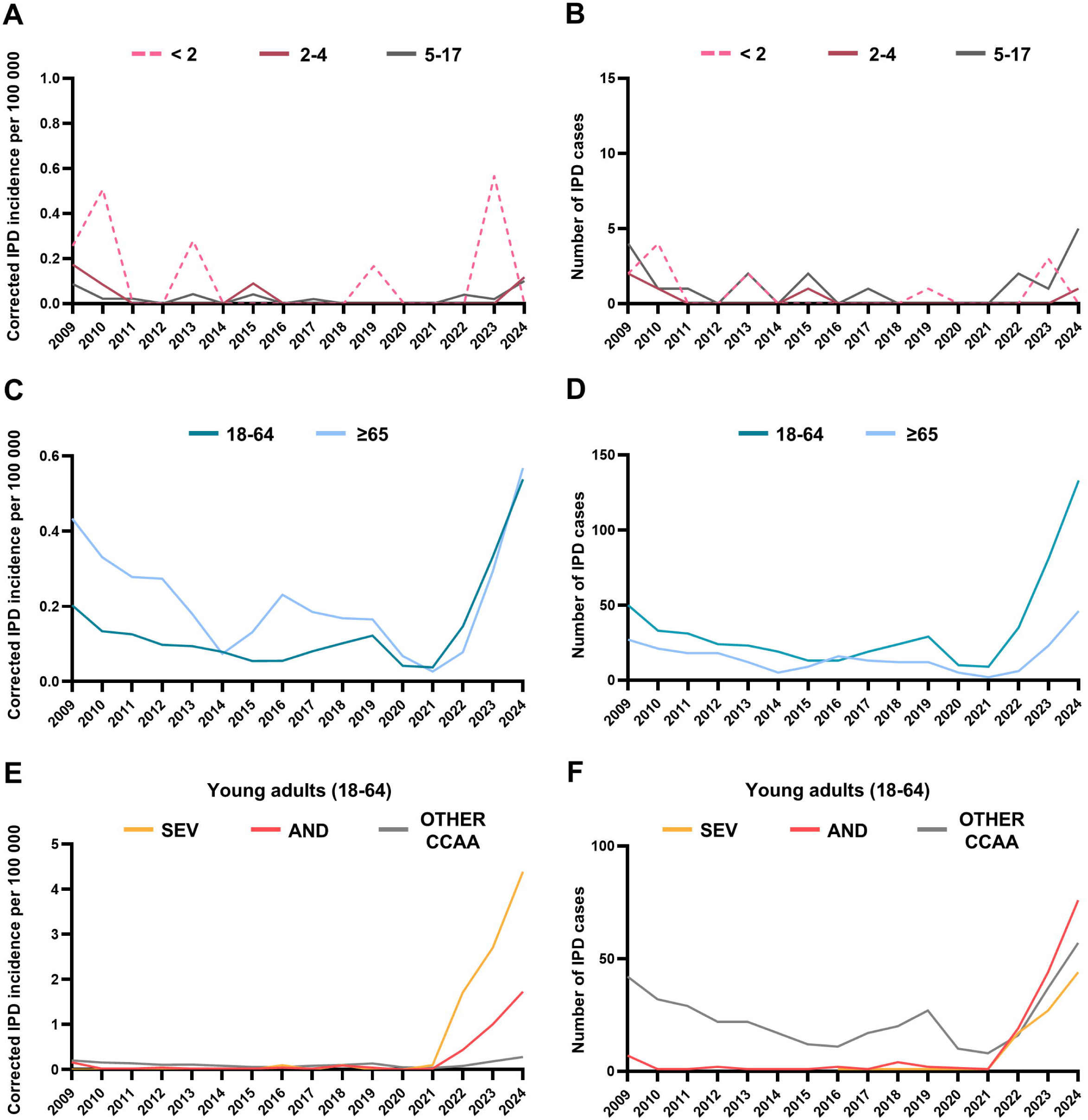
Corrected annual incidence and number of IPD cases of serotype 4 in Spain in pediatric and adult populations during 2009-24. IPD cases and incidence rates for the pediatric populations <2 years, 2–4 years and 0–17 years (A-B). IPD cases and incidence rates for adult populations 18-64 years old, and ≥ 65 years old (C-D). IPD cases and incidence rates for young adults (18-64 years) living in the subregion-povince of Seville (SEV), the region-CCAA of Andalusia (AND) or other Spanish regions (OTHER CCAA) (E-F).

In adults, the burden of IPD caused by serotype 4 declined with the introduction of PCV13 in children (Figure 1C-D) confirming herd-protection immunity against this serotype. During the middle vaccine period (2013 vs. 2009) incidence rates were significantly lower for serotype 4 (IRR, 0.25; 95% CI, 0.17–0.37 for the age group 18-64 years old and IRR, 0.36; 95% CI, 0.22–0.58 for adults ≥65 years old). These results were also confirmed when assessing the late PCV13 period (2017-19) vs. 2009 as pre-PCV13 year (Figure 1C-D and Table 1). Despite the low number of serotype 4 cases in the late vaccine period, there was a reduction of cases during COVID-19 (2020-21) due to the implementation of containment measures (Figure 1C-D and Table 1). Remarkably, after the end of containment measures, serotype 4 IPD cases spiked in all adult populations, but specifically in young adults (Figure 1C-D). The rise started in 2022 as found 9 cases in 2021, 35 cases in 2022, 81 cases in 2023 and 133 cases in 2024 (Figure 1D). This expansion of serotype 4 cases in young adults was significant when comparing the last three full epidemiological years (2022-24) versus all the other vaccine periods, even before the introduction of PCV13 (2022-24 vs. 2009: IRR, 1.63; 95% CI, 1.24-2.28 | 2022-24 vs. 2017-19: IRR, 3.36; 95% CI, 2.59-4.37, | 2022-24 vs. 2020-21: IRR, 8.57; 95% CI, 5.38-13.67, Figure 1C-D and Table 1). In the case of adults ≥65, this increase of serotype 4 cases after COVID-19 pandemic was also observed, being especially relevant in the last two full epidemiological years (2022-24 vs. 2020-2021: IRR, 6.77; 95% CI, 3.12-14.69, | 2022-24 vs. 2017-19: IRR, 1.83; 95% CI, 1.24-2.27; Figure 1C-D and Table 1).

During serotype 4 expansion in young adults in the last three years, we observed that most of these cases belonged to the same Spanish geographical CCAA/region (Andalusia) and specifically, the province/subregion of Seville (Figure 2). Therefore, we decided to study the number of cases and incidence per region-CCAA and province/subregion, observing that most of serotype 4 cases occurred in Andalusia, and specifically in Seville subregion (Figure 1E-F). In the last three years, 19 out of 35 cases in 2022, 44 out of 81 cases in 2023 and 76 out of 133 cases in 2024 appeared in Andalusia, and 57-89% of them were specifically in the province/subregion of Seville (Figure 1E-F and Figure 2). In terms of national dissemination, we found that in the year 2024, all different Spanish regions-CCAA notified IPD cases by serotype 4 in young adults although, the main accumulation of cases was still occurring in Seville and in other subregions of Andalusia (Figures 1C-D and Figure 2). In adults ≥65 we also observed a concentration of cases in Andalusia (40%, 30 out of 75 cases reported in Spain), of which 70% were reported in Seville (70%, 22 out of 30 cases reported in Andalusia).

**Figure 2.**
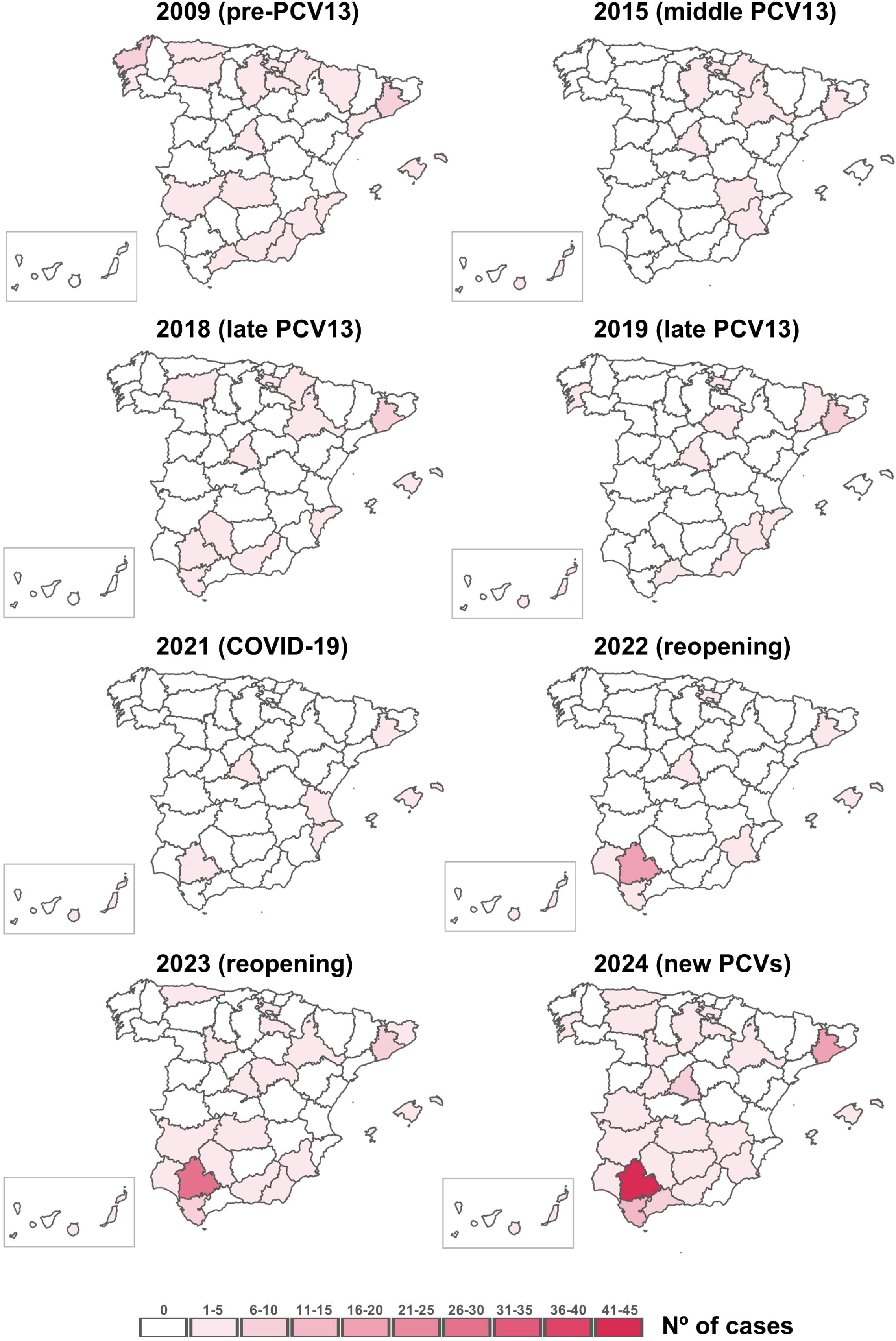
Geographic expansion of serotype 4 in young adults (18-64 years old) in Spain. The map shows the number of IPD cases notified to the SPRL by subregion and by epidemiological year (from January to December). Seville (SEV) is located in Andalusia which is in the south of Spain and is highlighted in the map.

### ST15063/CC801/GPSC162 dominates serotype 4 in recent expansion

We performed an in-deep WGS analysis of 397 serotype 4 clinical isolates from different vaccine periods: pre-PCV13 (n=51), middle PCV13 (n=15), late PCV13 (n=47), COVID-19 (n=5), and reopening including the last epidemiological year (n=279) (Supplemental Table S1). We sampled a strong representation of more than 60% of serotype 4 isolates for the period 2009-2024. Of all isolates tested, 17 belonged to the pediatric population, 58 to adults ≥65 years, and the majority to young adults (18-64 years old) as they had the highest burden of disease by serotype 4 (n=322) (Supplemental Table S1). Evaluating all vaccine periods, we found up to eleven different lineages causing IPD by serotype 4, being the four most common GPSC162 (n=271), GPSC27 (n=66), CC1866 (with not assigned GPSC) (n=17), and GPSC70 (n=14) (Figure 3A). In 2009, most isolates belonged to GPSC27 (56.9%; 29 out of 51 sequenced isolates) divided in ST205, ST247 and ST246 (Figure 3), but since PCV13 introduction, IPD cases by this lineage diminished. However, after COVID-19 pandemic there was a modest rise of cases by this lineage (Figure 3) but mostly due to ST247 (83.33%; 10 out of 12 GPSC27 isolates).

**Figure 3.**
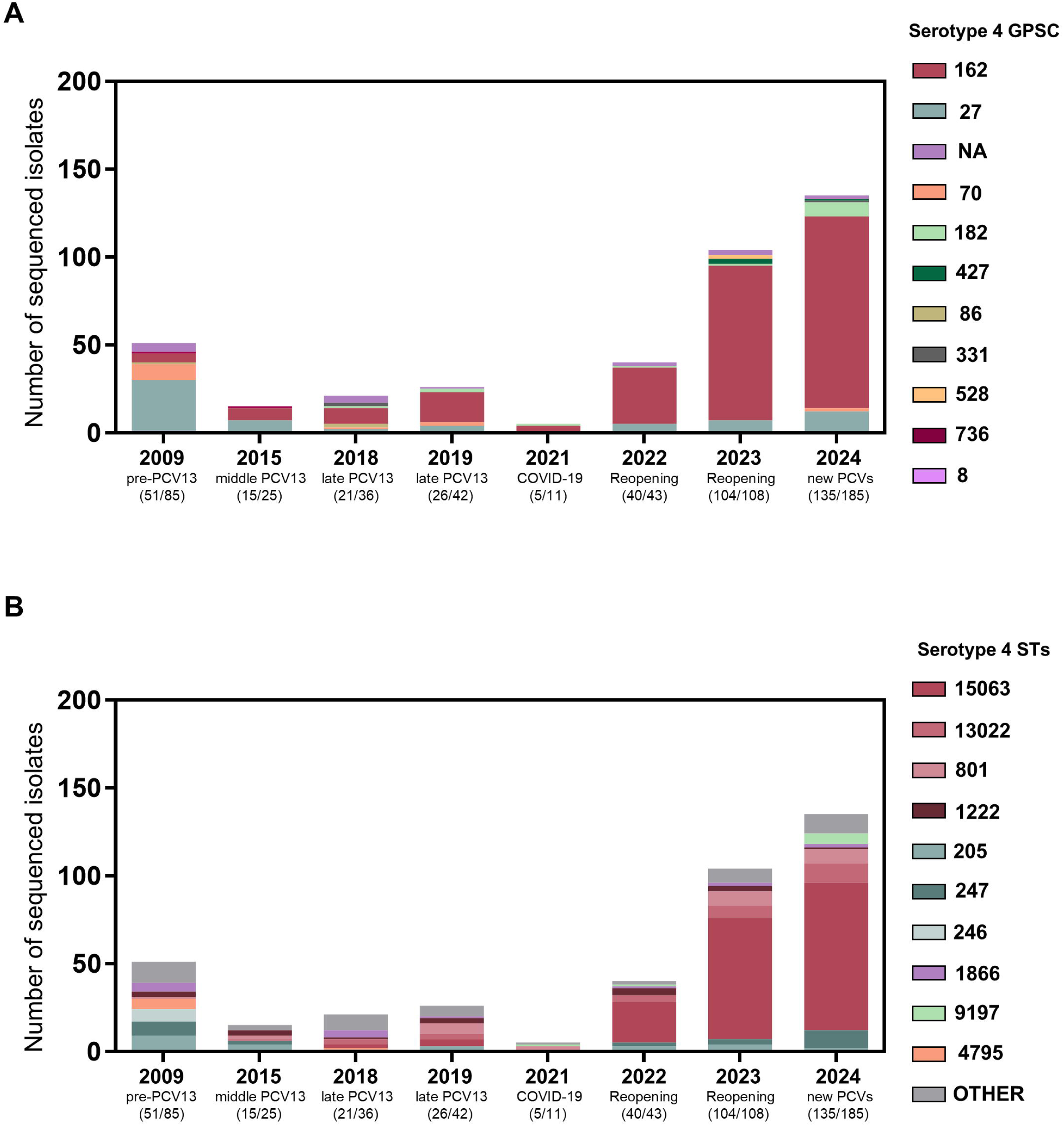
Trends in the distribution of lineages among serotype 4 isolates across different vaccine periods covering 2009-24. Serotype 4 GPSC distribution by year (A). Serotype 4 sequence-type (ST) distribution by year (B).

In contrast, we observed a remarkably evolution of GPSC162 associated with the rise of serotype 4, as there were very few cases by this lineage in the pre-PCV13 and middle vaccine periods. However, during the late PCV13 period, GPSC162 started to be the most frequent cause of IPD among serotype 4 although with a low burden of disease (Fig 3A). Nevertheless, after the uplift of containment measures, there was an outburst of serotype 4 cases in Spain mainly driven by ST15063 within GPSC162 (63%; 176 out of 279 sequenced isolates during 2022-24) (Figure 3). There were other STs producing IPD in the last three years in Spain, ST13022 (n=22) and ST801 (n=16), both also within GPSC162 (Figure 3). Moreover, most Andalusia isolates of serotype 4 from 2022-24 belonged to ST15063 (94.8%; 145 out of 153), and the same distribution was found for most Seville isolates (97.1%; 100 out of 103) (Figure 3 and Supplemental Table S1). An additional analysis evaluating all serotype 4 isolates and GPSC162 phylogenies from 2022-24 revealed that ≥ 97% of Andalusia isolates and all Seville isolates (100%) were closely related, being grouped within the ST15063 clade or a very closely related clade of the single *locus* variant ST801 (Figure 4). Both the dominant ST15063 and ST801 were more distantly related to other STs within GPSC162 such as ST1222 and ST13022, and most of these distantly related isolates originated from other Spanish regions (Figure 4 and Supplemental Figure S2). Genomic analysis of serotype 4 isolates from other Spanish regions were spread along the phylogenetic trees, but we observed a highly related cluster from Andalucia and specifically from Seville (3 to 15 nucleotide differences between Seville ST15063 isolates) that suggest clonal differences between Andalusia and other Spanish CCAA/regions (Figure 4). For example, since the expansion of serotype 4 in 2022-24, IPD cases by serotype 4 in other regions presented more lineage diversity (Figure 4 and Supplemental Figure S2). For instance, GPSC27 was also important in the other Spanish CCAA/regions (17.46%; 22 out of 126), and within GPSC162 (63.49%; 80 out of 126), isolates contained more variability of STs. ST15063 was still the most frequent (38.8%; 31 out of 80), followed by ST13022 (27.5%; 22 out of 80), ST801 (15%; 12 out 80) and ST1222 (10%; 8 out of 80).

**Figure 4.**
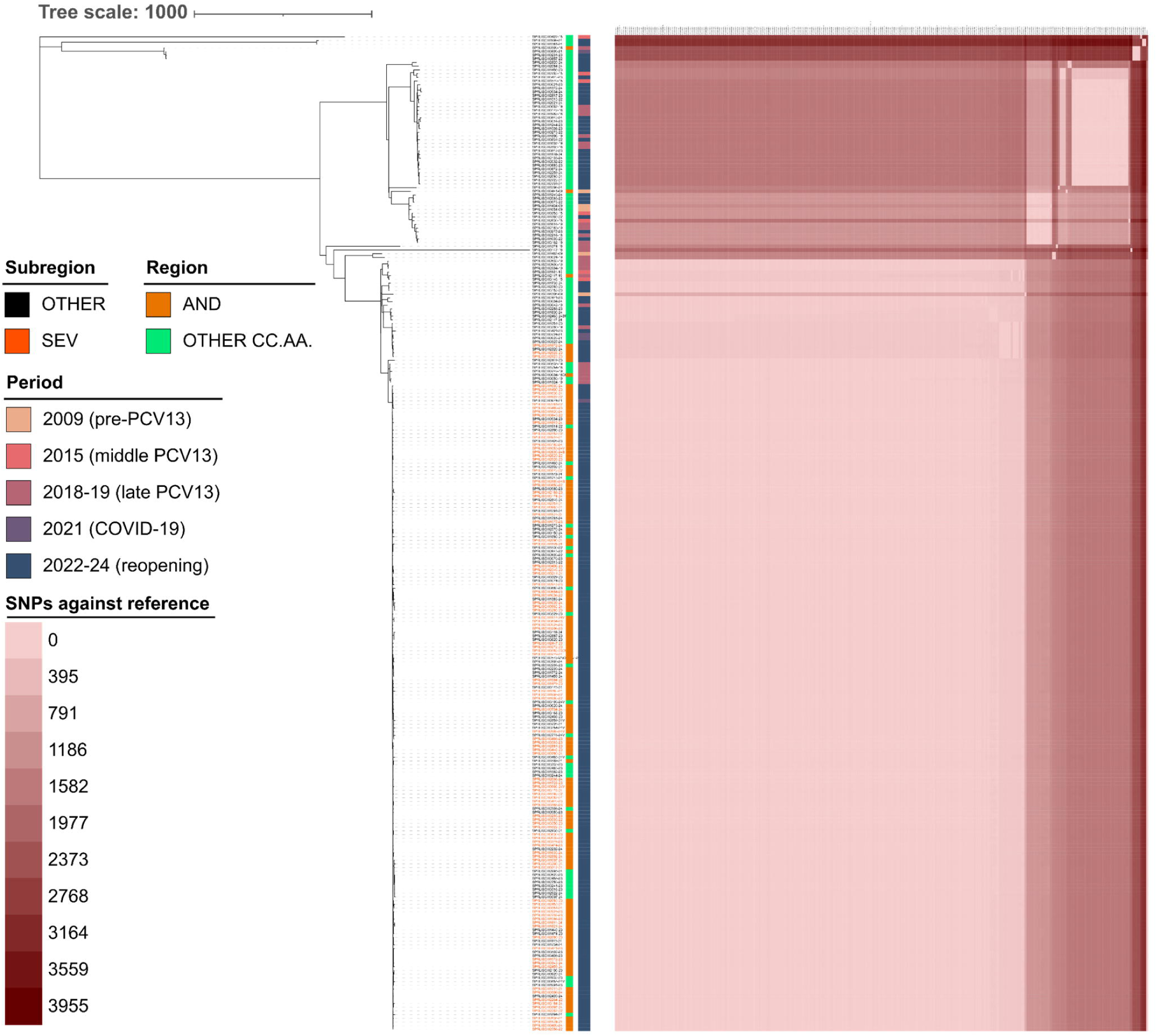
Phylogenetic tree of GPSC162 in Spain. The phylogenetic tree was obtained using the snippy, snippy-core, and Gubbins steps described in methods, and groups genomes into different clades based on the SNPs against the reference (GCA_09128732). Heatmap displaying differences in the SNPs, using GCA_09128732 as a reference. Geographic region was depicted (AND vs Other regions) and Seville isolates are highlighted in orange. Annotated data is described in the legend.

In general, the presence of mobile genetic elements (MGEs) was scarce in serotype 4 and is commented on supplemental results. Pangenome analysis revealed that the overall serotype 4 pangenome was very open, with nearly 40% of accessory genome (shell and cloud genes) (Figure 5A). Within serotype 4, GPSC162 and GPSC27 presented a higher total gene count (2202 and 2248 genes respectively) and a more open genome (19.08% and 20.14% of accessory genome respectively) compared to CC1866 (GPSC not assigned) (gene count of 1975 and 5.11% of accessory genome) and GPSC70 (gene count of 2033 and 10.28% of accessory genome) (Figure 5A). This also could be observed in the differences between total and conserved genes (Figure 5A). We investigated the pangenome relationship within GPSC162, observing that ST15063 presented a smaller total gene count (1999 genes) and very closed pangenome with only 5.5% of accessory genome, whereas other STs presented a higher total gene count (2153 genes) and 18.1% of accessory genome (Figure 5B).

**Figure 5.**
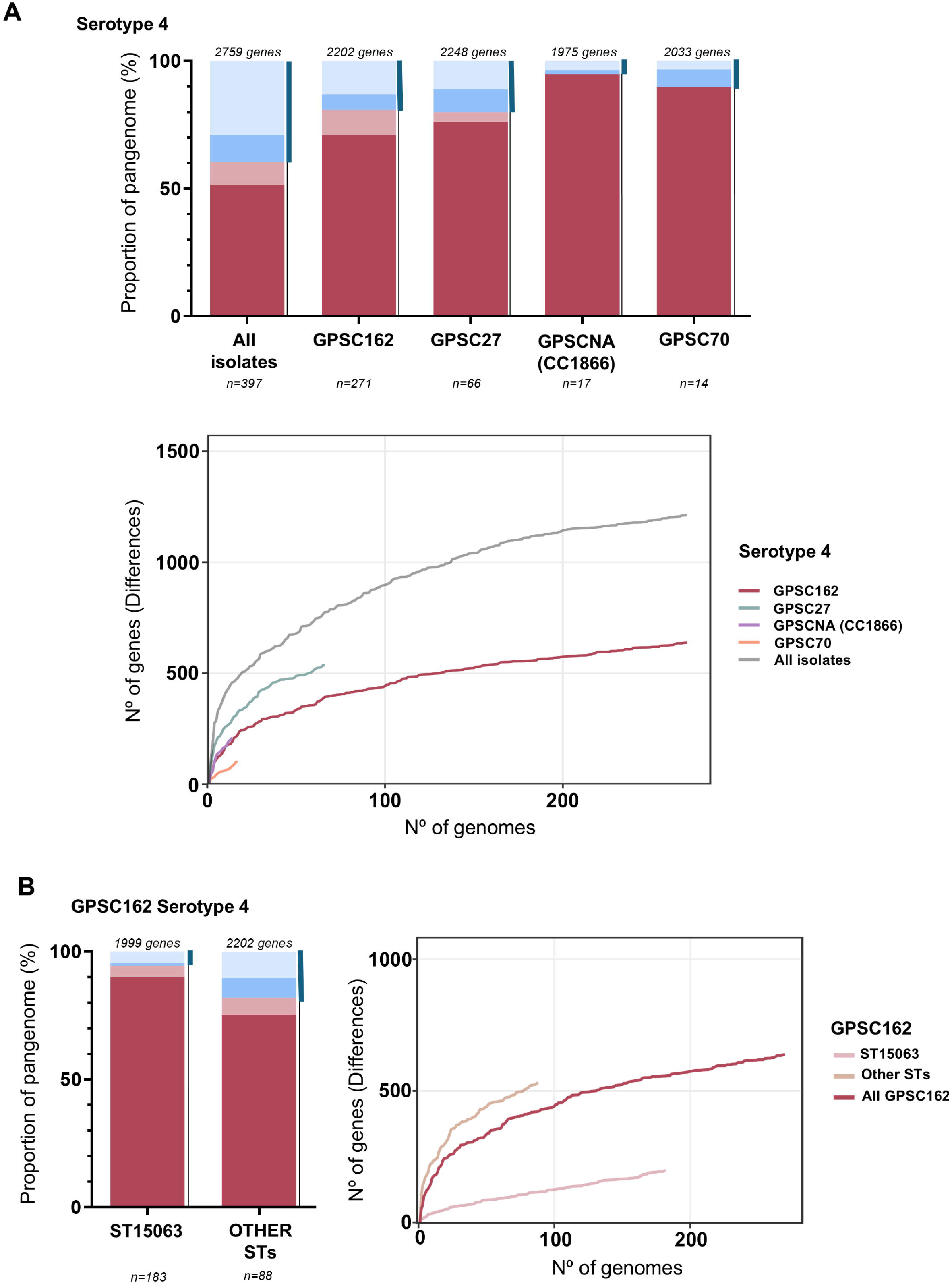
Analysis of the serotype 4 pangenome and major lineages. Percentage of core and accessory genome and relationship analysis between the total and conserved genes of overall serotype 4 isolates and the major lineages. The X-axis shows the number of genomes and the Y-axis the difference between the total and conserved genes in the pangenome analysis for each added genome (A). Percentage of core and accessory genome and relationship analysis between the total and conserved genes of overall GPSC162 isolates and ST15063. The X-axis shows the number of genomes and the Y-axis the difference between the total and conserved genes in the pangenome analysis for each added genome (B). Thick blue line highlights the proportion of the accessory genome (shell and cloud genes). GPSC not assigned (GPSCNA).

### Serotype 4 IPD patients in Seville are associated with substance use as a risk factor

Due to the localized expansion of serotype 4 cases in Seville, Andalusia, since the end of COVID-19 containment measures, we analyzed the clinical characteristics of young-adult patients with IPD in this subregion during 2022–2024. Table 2 summarizes the clinical and demographic characteristics of hospitalized patients with IPD cases by serotype 4 compared with those infected by other serotypes. The mean age was identical between groups, while the proportion of male patients was slightly higher in the serotype 4 group (*p* = 0.061). The increase of serotype 4 cases was not associated with different comorbidities such as COPD, asthma, other respiratory diseases, arterial hypertension, diabetes mellitus, and dyslipidemia (Table 2).

However, we observed significant differences when comparing living conditions and risk factors. Hospitalized patients with IPD by serotype 4 were more frequently homeless or imprisoned (9.4% vs. 1.5%, *p* = 0.017). In addition, we found significant associations of serotype 4 in patients with a record of risk factor for IPD including smoking (90.6% vs. 68.7%, *p* < 0.001), alcohol consumption (43.5% vs. 35.8%, *p* = 0.036), parenteral drug use (17.6% vs. 6.9%, *p* = 0.021), inhaled drugs such as cocaine and cannabis (37.6% vs. 16.0%, *p* < 0.001), and polydrug use (34.1% vs. 9.9%, *p* < 0.001) (Table 2).

Conversely, hospitalized patients with IPD by other serotypes were more frequently affected by comorbidities typically associated with increased IPD risk, such as immunosuppression of any cause (26.7% vs. 5.9%, p < 0.001) and chronic corticosteroid use (25.9% vs. 8.2%, *p* = 0.002). Patients hospitalized by other serotypes also showed a higher proportion of severe COVID-19 cases (7.6% vs. 0%, *p* = 0.007). Finally, vaccination status differed between groups: pneumococcal vaccination (PCV13 or PCV13 + PPV23) was more common in patients with IPD by other serotypes (25.2% vs. 8.2%, *p* = 0.008) (Table 2).

### Cigarette smoke exposure and H3N2 influenza virus infection increase infectious titers of S. pneumoniae serotype 4 in lung respiratory cells

We first evaluated the infectivity of different serotype 4 strains including clinical isolates from the Andalusia (Seville) outbreak (ST15063/CC801/GPSC162] or non-outbreak related [ST205/CC205/GPSC27]. Our results confirmed that serotype 4 strains of ST15063 had an increased ability to infect lung cells than strains of ST205 and this effect was even higher in the presence of CSE (Figures 6A-B). Co-infection experiments with influenza viruses confirmed that serotype 4 had increased infection rates of human lung cells in the presence of a human influenza H3N2 virus but not H1N1 and this effect was significantly higher with the combination ST15063/CC801/GPSC162 from the outbreak and influenza H3N2 virus (Figure 6C).

**Figure 6.**
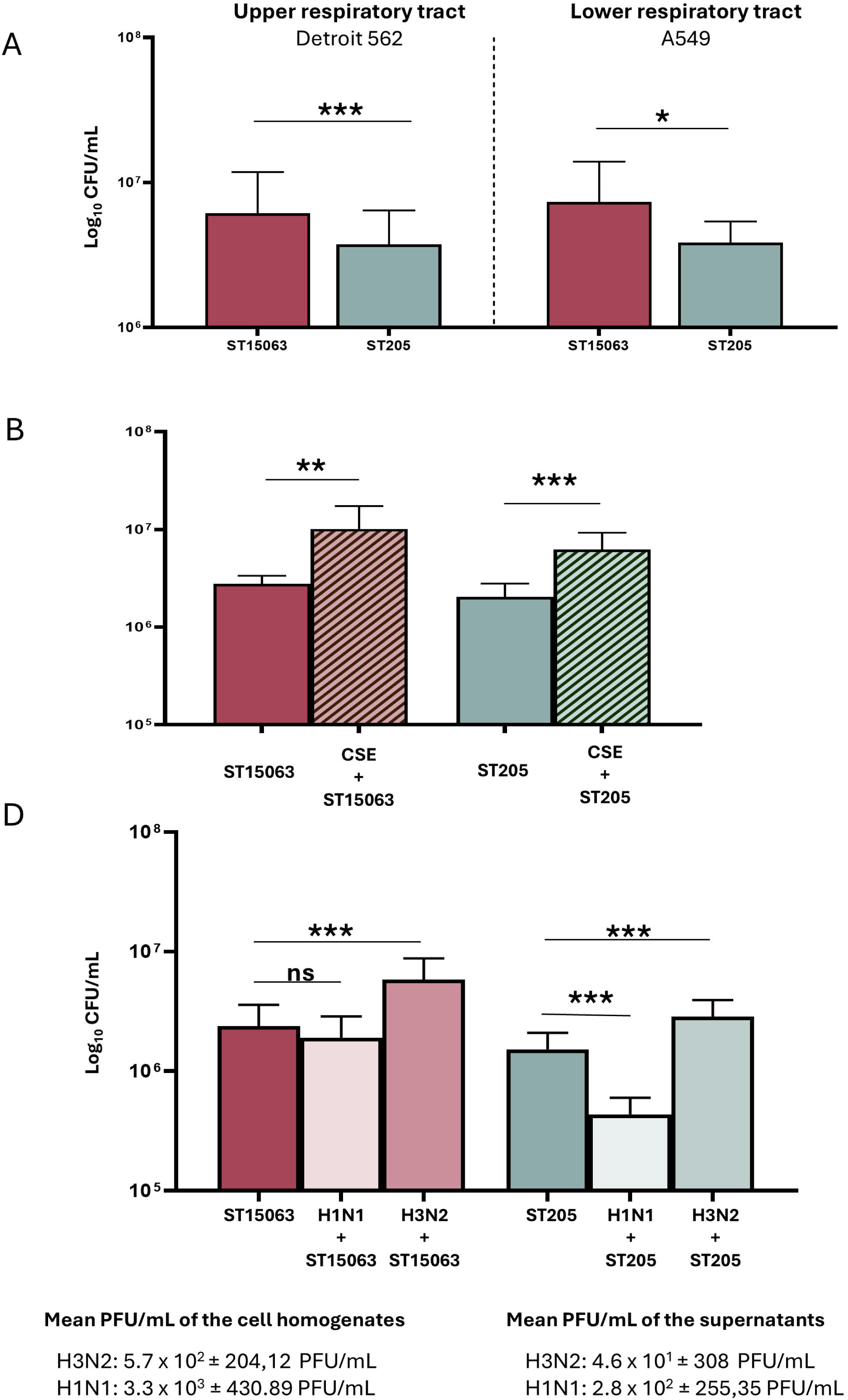
Comparative adhesion of serotype 4 isolates to human lung and nasopharyngeal epithelial cells: contribution of cigarette smoke and influenza infection in CC801 (ST15063, Andalusia outbreak) versus CC205 (ST205). (A) Adhesion to human nasopharyngeal (Detroit 562) and lung (A549) epithelial cells. (B) Effect of cigarette smoke exposure on adhesion to lung epithelial cells (A549). (C) Differences in the effect on secondary pneumococcal adhesion by serotype 4 (S4) following primary infection with human influenza A virus (H1N1 and H3N2 variants).

## Discussion

Our study documents the unexpected expansion of serotype 4 causing IPD in young adults in Spain after the COVID-19 pandemic and the rise to adults ≥65 years population, with most cases concentrated initially in the CCAA/region of Andalusia, and particularly in the province/subregion of Seville. Incidence by serotype 4 has been very low in Spanish children, and its incidence in adults of all age groups showed a declining trend after the introduction of PCV13 in pediatric populations [19,20]. Given the low vaccination coverage with PPV23 or PCV13 among Spanish adults [2,4], most of the reduction in IPD caused by PCV13 serotypes in both younger and older adults has been attributed to herd immunity [2,3].This specific resurgence of serotype 4 in adults is puzzling because pediatric PCV13 coverage has remained consistently high in Spain (>95%) [4], even during the pandemic, and IPD cases by serotype 4 IPD have not upturned in children. Furthermore, studies in the UK have shown that antibody responses to serotype 4 after PCV13 are robust, making it unlikely that the resurgence is due to reduced vaccine-induced herd immunity [6,21].

The unexpected rise of serotype 4 in young adults has also been observed in other countries. The UK reported an increase in serotype 4 from 23 cases in season 2019-20 to 131 cases in season 2022-23 and was also specific among young adults [6]. Moreover, Canada reported an upsurge of IPD cases after COVID-19, and they observed that serotype 4 was the most prevalent serotype in adults in 2022 (28.4%) [7]. Interestingly, the same region of Canada reported an increase of serotype 4 between 2010-2018, particularly among homeless persons, with phylogenetic evidence of both clonal clusters that are in line with outbreak dynamics and more genetically diverse strains disseminating in the community [22]. In addition, the USA has also reported increased proportions of IPD cases by serotype 4 in adults experiencing homelessness [8,9]. In northern Europe, similar outbreaks have been described, but in shipyard workers [10].

One of our main objectives was to decipher the cause of this specific expansion. From an epidemiological standpoint, our findings resemble outbreak dynamics previously described in USA and Canada [8,9,22], as we describe an upsurge of cases in a period of three years associated with a specific subregion in Spain. Outbreaks of IPD are rare events and when they occur, are often associated with vulnerable populations and groups living in crowded conditions, and a temporal relation is more difficult to establish as pneumococcal outbreaks may occur over a period of several years [22]. Our genomic analysis further supports this interpretation, as the expansion of serotype 4 cases in Seville was almost exclusively driven by ST15063, a lineage within GPSC162 closely related to ST801. Nearly all isolates from Seville formed a tight phylogenetic cluster (3 to 15 nucleotide differences between Seville ST15063 isolates), strongly suggesting clonal expansion rather than random reintroduction of diverse serotype 4 strains. Moreover, these strains presented a very closed pangenome indicating highly clonality. In contrast, isolates from other Spanish regions were genetically more diverse, encompassing multiple STs within GPSC162 and other lineages such as GPSC27 or GPSC182. This pattern suggests localized amplification of ST15063 in Seville, whereas other regions have a more heterogeneous molecular epidemiology.

Association of lineage GPSC162 with outbreaks in other countries has been previously shown although it was linked to ST801 [10]. In USA and Canada, serotype 4 outbreaks were produced by different STs, including ST244 and ST695 in both countries [9,22], ST10172 specifically in the USA and ST205 in Canada [8,9,22]. In Spain, ST244 and ST205 within GPSC27 were present and dominant in the pre-PCV13 period, although the number of cases was scarce after PCV13 introduction and could not be associated with specific populations and were geographically spread. In the last years, there were few cases by ST205, as ST247 predominated within GPSC27 after COVID-19, and most of isolates did not follow a geographical pattern. In England, despite not correlating serotype 4 cases with clinical data or with certain populations, GPSC162 has also displaced GPSC27 as the main serotype 4 lineage, and IPD cases were driven by ST801 and ST15063 [6].

The clinical profile of patients suffering IPD in Seville in the last three years provides additional insight. Compared to hospitalized patients with IPD by other serotypes, serotype 4 cases were disproportionately associated with substance use as risk factors (including tobacco, alcohol, parenteral drugs, inhaled drugs and polydrug consumption) and pneumonia was the primary diagnosis at hospital admission. This is consistent with the increased adhesion pattern to lung cells that was enhanced in the presence of CSE.

In contrast, comorbidities classically linked to IPD, such as immunosuppression by any cause or chronic corticosteroid use, were less frequent among serotype 4 patients. This pattern closely mirrors outbreaks in Canada and the United States, where serotype 4 disproportionately affected socially vulnerable young adults, often without underlying chronic disease but with high rates of substance use [8,9,22]. Specifically, in Canada, serotype 4 cases were also associated with illicit drug use, alcohol abuse, and smoking [22]. In the case of social vulnerability (we analyzed homelessness and imprisonment), we observed higher proportions in the serotype 4 IPD group (9.4%) than in IPD group by other serotypes (1.5%), although ratios were lower than those reported in Canada and USA, where approximately 47% and 36% of patients with IPD by serotype 4, respectively, experienced homelessness [8,9,22]. Nevertheless, despite lower proportion of social vulnerability than other outbreaks, it is markedly higher than the general Spanish population. In Spain, an estimated 23,000–30,000 people experience homelessness and around 55,000 are imprisoned, together representing only about 1.7% of the population (Instituto Nacional de Estadística, España). Taken together, these findings suggest that the expansion of serotype 4 in Seville represents a socio-behaviorally driven outbreak mainly affecting young adults from Seville, rather than waning pediatric herd immunity, facilitated by clonal spread of ST15063. This is a concerning situation because in the year 2024, the rise of serotype 4 started to expand to other Spanish regions, acquiring a national dissemination pattern, and is worrisome because in the last years, we also found an increase of cases affecting adults over 65 years old.

Another important factor increasing the susceptibility to pneumococcal disease, is the contribution of influenza virus infections [23]. Hence, serotype 4 strains showed increased infection rates in the presence of a human influenza H3N2 virus but not H1N1 and this effect was greater for the serotype 4 outbreak lineage. This might explain the increased rates of IPD by serotype 4 in elderly populations as they have higher incidence rates of influenza.

From a public health perspective, these findings stress the importance of targeted vaccination strategies - not only against pneumococcus but also against other respiratory viruses, such as influenza-, considering both direct immunization and adult-to-adult transmission. Despite the availability of new higher valent vaccines PCV15 and PCV20, both with serotype 4 in their formulations, adult vaccine uptake in Spain remains low [4]. Experiences from Canada indicate that homelessness and drug use should be considered explicit indications for pneumococcal vaccination, although practical challenges in reaching high vaccine coverage rates in these populations persist [22]. Given the strong association of serotype 4 with behavioral risk factors in Seville, tailored interventions, including vaccination campaigns targeting young adults with substance use, and enhanced outbreak surveillance, are warranted.

In summary, the post-pandemic expansion of serotype 4 IPD in Seville represents a clonal outbreak driven by ST15063 within GPSC162, affecting predominantly young adults with high rates of substance use. These findings parallel international experiences of serotype 4 outbreaks in substance users, homeless and occupational groups, stressing the need for better implementation of adult vaccination, and the need of ongoing genomic surveillance to detect and contain future pneumococcal outbreaks.

## Author contributions

JY is the scientific leader of the national reference laboratory and was responsible for the management of epidemiological surveillance data. JS and JY wrote the first draft of the paper. CPG, JL, MEAA, ML, RG, GL, FEA, VY, MS, JD, JMC, AGS, MD, JS and JY provided technical support for the study. CPG, MD, JL, JS and JY contributed to the study conception, design, data analysis, and interpretation. JS contributed to the WGS analysis and interpretation. All authors contributed to the review of the different drafts and approved all versions of the manuscript.

## Ethics statement

IPD cases and microbiological data are part of the national surveillance program of Invasive Pneumococcal Disease and do not require individual consent or Ethics Committee approval. All data analyzed is anonymized. Clinical and demographic data of young adults (18–65 years) diagnosed with IPD by any serotype between January 2022 and December 2024 from the five main hospitals in Seville were anonymized, and the study was approved by the Ethics Committee of the Hospital Universitario Virgen del Rocío (ENISE4, SICEIA-2025-002346).

## Data availability statement

All reads used for WGS analysis were deposited at the European Nucleotide Archive (Supplemental Table S1). All epidemiological and experimental data requests should be submitted to JS or JY. Requests will be assessed for scientific rigour before being granted, and a data-sharing agreement might be required.

