## Supplemental material for "Outburst of serotype 4 IPD after COVID-19 is driven by ST15063/GPSC162 lineage associated with high-risk behaviors and greater virulence linked to influenza H3N2 virus coinfection and cigarette smoke"

### Content

**Figure S1: Map of Spain divided into provinces/subregions……………………...2**

**Figure S2: Phylogenetic tree of serotype 4 in Spain…………………………………3**

**Supplemental methods……………………..………………………………………4**

**Supplemental results……..……………………..…………………………………7**

**Supplemental bibliography……..………………..…………………………………8**

**Table S1……………………..………………..……………………………………9**

**Table S2……………………..………………..………………………………28**

.

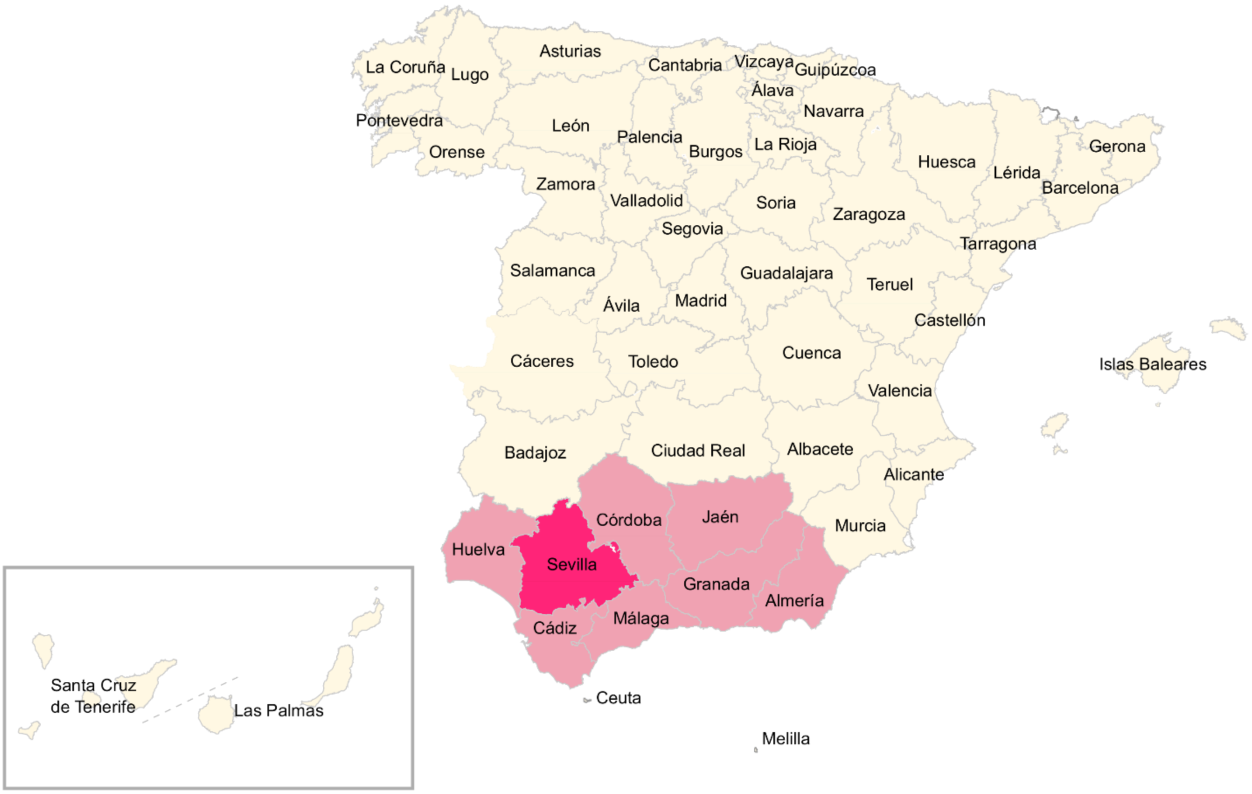

**Supplemental Figure S1. Map of Spain divided into provinces/subregions.** The CCAA/region of Andalusia (AND) is highlighted in pink, and the province/subregion of Seville (SEV) within AND is highlighted in bright pink.

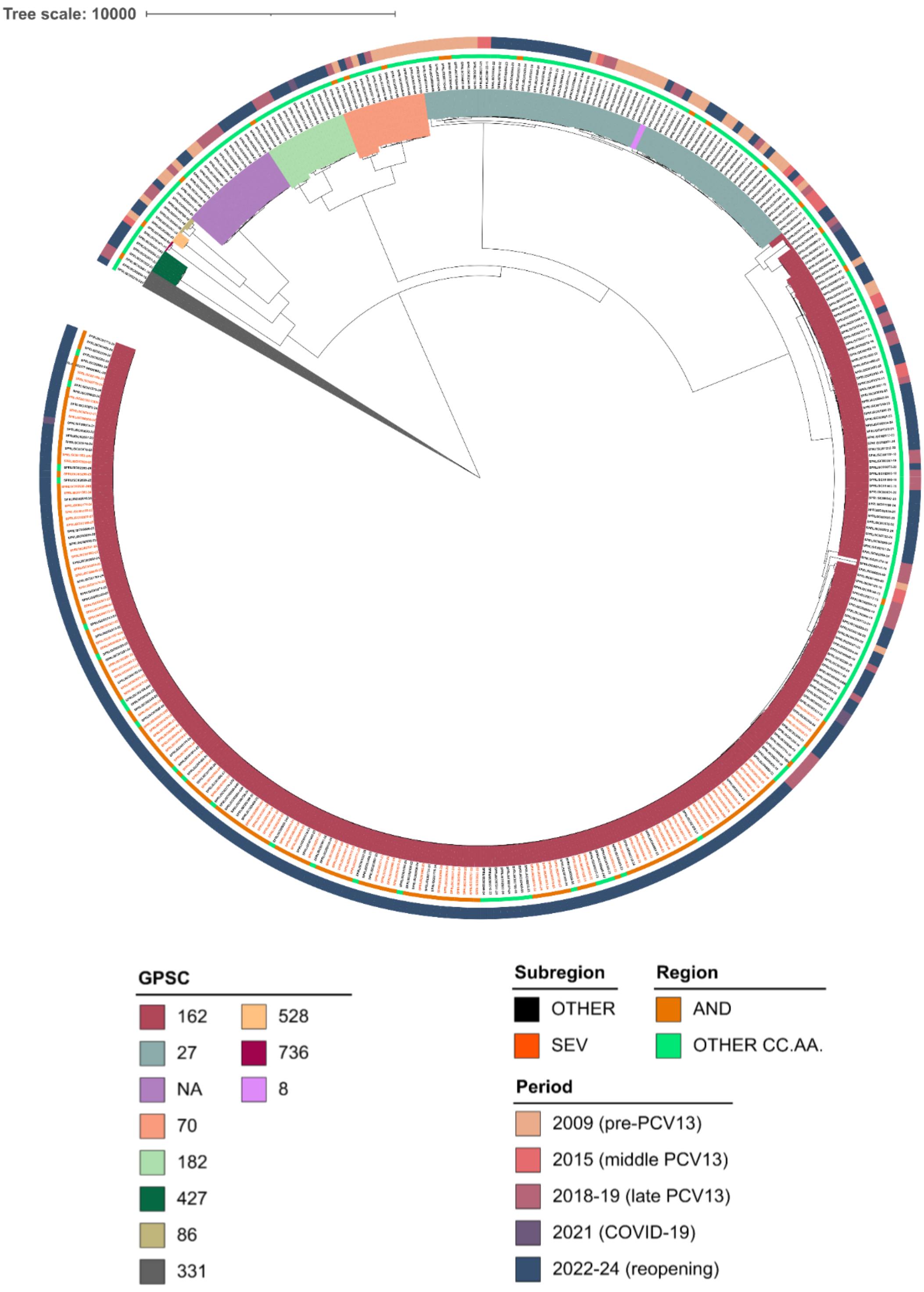

**Supplemental Figure S2. Phylogenetic tree of serotype 4 in Spain.** The phylogenetic tree was obtained using the snippy, snippy-core, and Gubbins steps described in methods, and groups genomes into different clades based on the SNPs against the reference (GCA_09128732). The geographic region was depicted (AND vs Other Spanish regions), and Seville isolates are highlighted in orange. Annotated data is described in the legend.

### SUPPLEMENTAL METHODS

*Bioinformatic analysis*

WGS pipeline included quality assessment with FASTQC (https://www.bioinformatics.babraham.ac.uk/projects/fastqc/), removal of low-quality reads, decontamination from non-target bacterial sequences, and de novo genome assembly using Shovill (https://github.com/tseemann/shovill). Bactopia integrates multilocus sequence typing (MLST; https://github.com/tseemann/mlst) and genome annotation with Prokka (https://github.com/tseemann/prokka) without additional processing. Pneumococcus-specific analyses were performed with PBP-typer (https://github.com/rpetit3/pbptyper). Preliminary screening for antimicrobial resistance (AMR) genes and virulence factors was carried out using Abricate (https://github.com/tseemann/abricate). For phylogenetic analysis, single-nucleotide polymorphism (SNP) calling was performed with Snippy (https://github.com/tseemann/snippy), phylogenetic trees were inferred using RAxML-NG (https://github.com/amkozlov/raxml-ng), and recombination events were identified with Gubbins (https://github.com/nickjcroucher/gubbins)

*Nasopharyngeal adhesion assays using Detroit 562 cells*

To evaluate differences in nasopharyngeal adhesion among distinct serotype 4 sequence types (STs), Detroit 562 cells (ATCC CCL-138) were used. Cells were maintained in RPMI 1640 supplemented with 10% heat-inactivated fetal bovine serum (56°C for 30 min), sodium pyruvate, L-glutamine, and penicillin/streptomycin, and incubated at 37°C in a humidified atmosphere containing 5% $CO_2$.

Two days prior to infection, cells were detached using trypsin–EDTA, washed, and seeded at $2 \times 10^5$ cells per well in 24-well plates to reach approximately 95% confluence on the day of the experiment (doubling time ≈88 h).

On the day of the assay, monolayers were washed three times with 1× PBS and 1 mL of antibiotic-free RPMI 1640 was added per well. Cells were infected with $7.5 \times 10^6$ CFU of *S. pneumoniae* in 50 µL to achieve a multiplicity of infection (MOI) of 25 bacteria per cell. Plates were incubated for 1 h at 37°C with 5% $CO_2$ to allow bacterial adhesion.

Following incubation, supernatants were aspirated and wells were washed three times with 1× PBS to remove non-adherent bacteria. Cells were lysed with 200 µL of 0.025%

saponin and incubated at room temperature for 10 min. Lysates were homogenized, serially diluted (10-fold), and plated on blood agar to determine adhered CFU/mL the following day. Experiments were performed in at least three independent biological replicates.

*Assessment of S. pneumoniae adhesion to A549 lung epithelial cells with or without cigarette smoke extract (CSE)*

CSE was obtained from research cigarettes (nicotine 0.73 mg, tar 9.4 mg, CO 12 mg which is equivalent with the content of commercial cigarettes) These cigarettes were supplied by the University of Kentucky. Briefly, one research cigarette was combusted using a 50 ml syringe-modified dispositive connected to a sterile gas wash bottle through a three-way stopcock as previously described [1]. This system draws the cigarette smoke into the sterile gas wash bottle containing 5 ml of tissue culture medium (RPMI 1640) and the procedure drawing the smoke into the bottle was repeated 5 times with vigorous manual shaking of the gas wash bottle to expose the RPMI medium to the smoke. Combustion of cigarettes was repeated using 8 different research cigarettes and a final volume of 50 ml of RPMI (proportion of 1 cigarette each 5 ml of RPMI) and the cigarette smoke solution was frozen in aliquots at -80°C for further use. This liquid solution was stored as CSE 100%.

Experiments assessing pneumococcal adhesion to the pulmonary epithelium were conducted using A549 human lung epithelial cells (CCL-185; ATCC). Cells were cultured in RPMI 1640 medium containing phenol red and L-glutamine, supplemented with 10% heat-inactivated fetal bovine serum (FBS) and 5% penicillin–streptomycin, and incubated at 37°C in a humidified atmosphere containing 5% $CO_2$.

Cells were seeded in 24-well plates at a density of $2 \times 10^5$ cells per well one day prior to the experiment, so that by the following day they reached near-confluence.

For infection, $5 \times 10^6$ CFU/mL of *S. pneumoniae* were added to each well, corresponding to a multiplicity of infection (MOI) of 25 bacteria per cell. Plates were incubated for 1 h at 37°C with 5% $CO_2$ to allow bacterial adhesion to the lung epithelial monolayer.

Following incubation, non-adherent bacteria were removed by washing the monolayers three times with 1× PBS. Cells were lysed with 200 µL of 0.025% PBS–saponin for 10 min at 37°C, and lysates were homogenized. Serial 10-fold dilutions of the lysates were plated on blood agar to determine the number of adherent bacteria (CFU/mL). All experiments were performed in at least three independent biological replicates.

To evaluate the effect of cigarette smoke extract (CSE) on pneumococcal adhesion, A549 cells were pre-exposed to 5% CSE for 24 h prior to bacterial infection. The following day, cells were washed twice with 1× PBS, and infection with serotype 4 strains was performed as described above.

*Interactions between human influenza variants and pneumococcal serotype 4 STs*

To carry out co-infections between influenza and *Streptococcus pneumoniae*, the A549 cell line was used. In order to determine an appropriate influenza multiplicity of infection (MOI) for each viral strain employed, *influenza A/New Caledonia/20/1999* (H1N1 subtype) or *influenza A/Darwin/9/2021* (H3N2 subtype), growth curves were performed at 4, 6, 8, and 24 hours in A549 cells at MOIs of 0.1 and 0.5 in infection medium for each virus (data not shown). An MOI of 0.5 was selected for both influenza strains to achieve comparable levels of infection across experiments, ensuring reproducibility in co-infection assays.

Experiments evaluating co-infection with influenza viruses were conducted using A549 cells cultured in DMEM supplemented with 0.2% BSA, 1 mM $MgCl_2$, and 0.9 mM $CaCl_2$. Briefly, $2\times10^5$ cells were seeded the day before and cultured at 37 °C with 5% $CO_2$. The following day, when cells reached approximately 95% confluence, they were infected with *influenza A/New Caledonia/20/1999* or *influenza A/Darwin/9/2021* at an MOI of 0.5 in infection medium. After 1 hour of viral adsorption, the medium was removed, and cells were incubated with 500 µL of supplemented DMEM for 24 hours at 37 °C with 5% $CO_2$.

The next day, cells were infected with *S. pneumoniae* (strains 582 [ST15063/CC801/GPSC162] or 724 [ST205/CC205/GPSC27]) at an MOI of 10. After 1 hour of bacterial adhesion, the supernatant was collected, and the monolayer was washed three times with PBS, as performed in the other assays. Cells were then homogenized in PBS to allow the homogenate to be used for both pneumococcal viable counts (CFU/mL) and influenza titration (PFU/mL). An aliquot of the homogenate was immediately diluted 1:10 to prepare the dilution series for CFU determination. Both the collected supernatant and the remaining homogenate were stored at −80 °C, and PFU titration was performed on a subsequent day.

The following day, cells were infected with *S. pneumoniae* (strains 582 [ST15063/CC801/GPSC162] or 724 [ST205/CC205/GPSC27]) at an MOI of 10. After 1 hour of bacterial adhesion, the supernatant was collected, and the monolayer was washed three times with PBS, as performed in previous assays. Cells were then homogenized in 200 µL of PBS, enabling the homogenate to be used for both pneumococcal viable counts (CFU/mL) and influenza titration (PFU/mL). An aliquot of the homogenate was immediately diluted 1:10 to prepare the dilution series for CFU determination. The collected supernatant and the remaining homogenate were stored at −80 °C, and PFU titration was performed on a subsequent day.

To quantify PFU/mL in both the supernatant and the cell homogenate, collected samples were first thawed and serially diluted in 1× PBS. 12-well plates, seeded the previous day with $3.5 \times 10^5$ MDCK cells per well, were washed once with 1 mL of 1× PBS. 250 µL of each sample dilution was added to the wells and incubated for 1 h at room temperature. After washing, 1 mL per well of a 2% agar overlay was added, allowed to solidify for 15 min at room temperature, and the plates were incubated at 37°C with 5% $CO_2$ for 48 h.

Cells were then fixed overnight at 4°C with 1 mL/well of 4% formalin. After washing with PBS-T (1× PBS + 0.05% Tween 20), monolayers were immunostained with 200 µL/well of polyclonal mouse serum in blocking buffer (5% milk in 1× PBS) for 1 h on a plate rocker. Following a single PBS-T wash, 200 µL/well of HRP-conjugated goat anti-mouse IgG Fc secondary antibody in blocking buffer was added for 1 h at room temperature with gentle rocking. Plates were washed again, and 150 µL/well of KPL TrueBlue substrate was applied for 15 min at room temperature. After a final wash, plaques were counted to determine viral titers [2].

### SUPPLEMENTAL RESULTS

#### *ST15063/CC801/GPSC162 dominates serotype 4 in recent expansion*

In general, the presence of mobile genetic elements (MGEs) was scarce in serotype 4 (Supplemental Table S1). Only 27 isolates showed the presence of MGEs related to resistance determinants leading to macrolides resistance [*erm*(B), *mef*(A) and *msr*(D)], cloranfenicol resistance (*cat*) and tetracycline resistance [*tet*(M)] (Supplemental Table 1). Most cases with MGEs were related to the presence of Tn*5252* family transposon (17 out of the 27 isolates) and belonged to GPSC162, only one present in ST15063 (Supplemental Table S1). The presence of prophages was also rare (n=13), with the majority belonging to PPH010 family of PPHs (Supplemental Table S1). Curiously, one of the isolates presented a non-conventional streptococcal phage (Javan340-like) with a phagic *lytA* uncommon for PPHs (Supplemental Table S1). PPHs were found throughout the different GPSCs. In addition, we found differences between lineages in the presence of pilus type I, as none of the GPSC162 or CC1866 (GPSC not assigned) isolates contained the pilus islet 1 (PI-1) encoding pilus type 1. On the contrary, all GPSC27 isolates, all GPSC70 isolates, and most GPSC182 isolates (92.9%, 13 out of 14) had the PI-1 (Supplemental Table S1).

**Supplemental Table 1. Data, accession numbers, and antibiotic/virulence factor analysis of clinical isolates sequenced in the study.** SEV, Seville; ND, non-detected by ICE berg 3.0; PI-1, pilus islet 1. [a]GENOME name is an internal code use for ISCIII sequencing and is not related to patient IDs.

| | METADATA | | | | | GENOTYPE | | | | | | | | | | AMR | | | | | | | MOBILE GENETIC ELEMENTS | VIRULENCE |
|---|---|---|---|---|---|---|---|---|---|---|---|---|---|---|---|---|---|---|---|---|---|---|---|---|
| GENOME[a] | GENOME ACCESION | YEAR | SEX | CCAA | SEVILLE | GPSC | CC | ST | aroE | gdh | gki | recP | spi | xpt | ddl | PBPTYPE | *cat* | *erm*(B) | *mef*(A) | *msr*(D) | *tet*(M) | Transposon | Pneumococcal prophage (PPH) | Pilus islet 1 (*rrgA*, *rrgB*, *rrgC*, *srtC-B*, *srtC-C*, *srtC-D*) |
| SPRLISCIII0004-18DU | ERS27214686 | 2018 | MAN | AND | | 162 | 801 | 15063 | 8 | 70 | 4 | 5 | 6 | 116 | 6 | 1_0_2 | . | . | . | . | . | | | |
| SPRLISCIII0020-24 | ERS27214687 | 2024 | MAN | AND | | 162 | 801 | 15063 | 8 | 70 | 4 | 5 | 6 | 116 | 6 | 1_0_2 | . | . | . | . | . | | | |
| SPRLISCIII0022-18 | ERS27214688 | 2018 | MAN | VAL | | NA | 1866 | 1866 | 1 | 11 | 1 | 15 | 103 | 1 | 70 | 0_0_0 | . | . | . | . | . | | | |
| SPRLISCIII0024-19 | ERS27214689 | 2019 | WOMAN | C-L | | 162 | 801 | 2361 | 8 | 8 | 4 | 1 | 6 | 116 | 6 | 23_0_2 | . | . | . | . | . | | | |
| SPRLISCIII0059-09 | ERS27214690 | 2009 | MAN | MUR | | 27 | 899 | 247 | 16 | 13 | 4 | 5 | 6 | 10 | 14 | 0_0_3 | . | . | . | . | . | | | PI-1 |
| SPRLISCIII0067-19 | ERS27214691 | 2019 | MAN | CAT | | 162 | 801 | 13022 | 8 | 32 | 4 | 1 | 6 | 4 | 6 | NEW_0_2 | . | . | . | . | . | | | |
| SPRLISCIII0102-09 | ERS27214692 | 2009 | MAN | NAV | | 27 | 899 | 246 | 16 | 13 | 4 | 5 | 6 | 10 | 18 | 0_0_3 | . | . | . | . | . | | | PI-1 |
| SPRLISCIII0114-24 | ERS27214693 | 2024 | MAN | MAD | | 27 | 899 | 247 | 16 | 13 | 4 | 5 | 6 | 10 | 14 | 0_0_3 | . | . | . | . | . | | | PI-1 |
| SPRLISCIII0122-15 | ERS27214694 | 2015 | MAN | C-L | | 27 | 899 | 247 | 16 | 13 | 4 | 5 | 6 | 10 | 14 | 0_0_3 | . | . | . | . | . | | | PI-1 |
| SPRLISCIII0143-09 | ERS27214695 | 2009 | MAN | GAL | | 27 | 205 | 205 | 10 | 5 | 4 | 5 | 13 | 10 | 18 | 0_0_3 | . | . | . | . | . | | PPH010 | PI-1 |
| SPRLISCIII0147-18 | ERS27214696 | 2018 | MAN | CAT | | 162 | 801 | 800 | 8 | 41 | 4 | 1 | 6 | 116 | 6 | 23_0_2 | . | . | . | . | . | | | |
| SPRLISCIII0150-24 | ERS27214697 | 2024 | MAN | AND | | 162 | 801 | 15063 | 8 | 70 | 4 | 5 | 6 | 116 | 6 | 1_0_2 | . | . | . | . | . | | | |
| SPRLISCIII0175-24 | ERS27214698 | 2024 | MAN | AND | | 162 | 801 | 15063 | 8 | 70 | 4 | 5 | 6 | 116 | 6 | 1_0_2 | . | . | . | . | . | | | |
| SPRLISCIII0177-18 | ERS27214699 | 2018 | MAN | CAT | | 162 | 801 | 13022 | 8 | 32 | 4 | 1 | 6 | 4 | 6 | NEW_0_2 | . | . | . | . | . | | | |
| SPRLISCIII0186-19 | ERS27214700 | 2019 | MAN | CAT | | 70 | 259 | 7026 | 24 | 5 | 1 | 1 | 15 | 12 | 43 | 2_44_NEW | . | . | . | . | . | | | PI-1 |
| SPRLISCIII0231-05 | ERS27214701 | 2015 | WOMAN | MAD | | 27 | 899 | 247 | 16 | 13 | 4 | 5 | 6 | 10 | 14 | 0_0_3 | . | . | . | . | . | | | PI-1 |

| | | | | | | | | | | | | | | | | | | | | | | | |
|---|---|---|---|---|---|---|---|---|---|---|---|---|---|---|---|---|---|---|---|---|---|---|---|
| SPRLISCIII0231-23 | ERS27214702 | 2023 | MAN | BAL | | 162 | 801 | 14136 | 8 | 12 | 4 | 1 | 17 | 116 | 72 | 23_0_2 | . | . | . | . | . | | |
| SPRLISCIII0238-09 | ERS27214703 | 2009 | WOMAN | RIO | | 27 | 899 | 246 | 16 | 13 | 4 | 5 | 6 | 10 | 18 | 0_0_3 | . | . | . | . | . | | PI-1 |
| SPRLISCIII0241-24 | ERS27214704 | 2024 | MAN | AND | SEV | 162 | 801 | 15063 | 8 | 70 | 4 | 5 | 6 | 116 | 6 | 1_0_2 | . | . | . | . | . | | |
| SPRLISCIII0260-24 | ERS27214705 | 2024 | MAN | VAS | | 182 | 2941 | 9197 | 97 | 5 | 1 | 226 | 36 | 1 | 14 | 2_4_2 | . | . | . | . | . | | PI-1 |
| SPRLISCIII0272-23 | ERS27214706 | 2023 | MAN | AND | SEV | 162 | 801 | 15063 | 8 | 70 | 4 | 5 | 6 | 116 | 6 | 1_0_2 | . | . | . | . | . | | |
| SPRLISCIII0273-22 | ERS27214707 | 2022 | WOMAN | CAT | | 162 | 801 | 13022 | 8 | 32 | 4 | 1 | 6 | 4 | 6 | NEW_0_2 | . | . | . | . | . | | |
| SPRLISCIII0277-23 | ERS27214708 | 2023 | MAN | RIO | | 162 | 801 | 1222 | 8 | 32 | 4 | 1 | 6 | 116 | 6 | 23_40_2 | . | . | . | . | . | | |
| SPRLISCIII0278-18 | ERS27214709 | 2018 | MAN | NAV | | 27 | 244 | 244 | 16 | 2 | 4 | 1 | 6 | 10 | 18 | 0_0_3 | *cat*(pC194)_1 | *erm*(B)_18 | . | . | tet(M)_8 | Tn*916*-family | PI-1 |
| SPRLISCIII0327-24 | ERS27214710 | 2024 | MAN | MAD | | 162 | 801 | 801 | 8 | 70 | 4 | 1 | 6 | 116 | 6 | 1_0_2 | . | . | *mef*(A)_2 | *msr*(D)_2 | . | ND | |
| SPRLISCIII0329-23 | ERS27214711 | 2023 | MAN | AND | | 162 | 801 | 15063 | 8 | 70 | 4 | 5 | 6 | 116 | 6 | 1_0_2 | . | . | . | . | . | | |
| SPRLISCIII0343-24 | ERS27214712 | 2024 | MAN | AND | SEV | 162 | 801 | 15063 | 8 | 70 | 4 | 5 | 6 | 116 | 6 | 1_0_2 | . | . | . | . | . | | |
| SPRLISCIII0347-24 | ERS27214713 | 2024 | WOMAN | AND | SEV | 162 | 801 | 15063 | 8 | 70 | 4 | 5 | 6 | 116 | 6 | 1_0_2 | . | . | . | . | . | | |
| SPRLISCIII0351-24 | ERS27214714 | 2024 | MAN | AND | SEV | 162 | 801 | 15063 | 8 | 70 | 4 | 5 | 6 | 116 | 6 | 1_0_2 | . | . | . | . | . | | |
| SPRLISCIII0358-15 | ERS27214715 | 2015 | MAN | VAS | | 162 | 801 | 1222 | 8 | 32 | 4 | 1 | 6 | 116 | 6 | 23_40_2 | . | . | . | . | . | | |
| SPRLISCIII0361-22 | ERS27214716 | 2022 | WOMAN | MAD | | 27 | 899 | 247 | 16 | 13 | 4 | 5 | 6 | 10 | 14 | 0_0_3 | . | . | . | . | . | | PI-1 |
| SPRLISCIII0396-24 | ERS27214717 | 2024 | MAN | AND | SEV | 162 | 801 | 15063 | 8 | 70 | 4 | 5 | 6 | 116 | 6 | 1_0_2 | . | . | . | . | . | | |
| SPRLISCIII0397-24 | ERS27214718 | 2024 | MAN | AND | SEV | 162 | 801 | 15063 | 8 | 70 | 4 | 5 | 6 | 116 | 6 | 1_0_2 | . | . | . | . | . | | |
| SPRLISCIII0399-18 | ERS27214719 | 2018 | MAN | AND | | 162 | 801 | 14136 | 8 | 12 | 4 | 1 | 17 | 116 | 72 | 23_0_2 | . | . | . | . | . | | |
| SPRLISCIII0406-23 | ERS27214720 | 2023 | WOMAN | AND | SEV | 162 | 801 | 15063 | 8 | 70 | 4 | 5 | 6 | 116 | 6 | 1_0_2 | . | . | . | . | . | | |
| SPRLISCIII0409-24 | ERS27214721 | 2024 | MAN | AND | SEV | 162 | 801 | 15063 | 8 | 70 | 4 | 5 | 6 | 116 | 6 | 1_0_2 | . | . | . | . | . | | |
| SPRLISCIII0427-15 | ERS27214722 | 2015 | WOMAN | CAN | | 162 | 1022 | 1022 | 8 | 8 | 4 | 1 | 15 | 1 | 20 | 23_0_2 | . | . | . | . | *tet*(M)_1 | Tn*916*-family | |
| SPRLISCIII0441-09 | ERS27214723 | 2009 | MAN | AND | | 162 | 801 | 1222 | 8 | 32 | 4 | 1 | 6 | 116 | 6 | 23_0_2 | . | . | . | . | . | | |

| | | | | | | | | | | | | | | | | | | | | | | | | |
|---|---|---|---|---|---|---|---|---|---|---|---|---|---|---|---|---|---|---|---|---|---|---|---|---|
| SPRLISCIII0485-24V | ERS27214724 | 2024 | WOMAN | C-M | | 162 | 801 | 15063 | 8 | 70 | 4 | 5 | 6 | 116 | 6 | 1_0_2 | . | . | . | . | . | | | |
| SPRLISCIII0503-22 | ERS27214725 | 2022 | MAN | AND | SEV | 162 | 801 | 15063 | 8 | 70 | 4 | 5 | 6 | 116 | 6 | 1_0_2 | . | . | . | . | . | | | |
| SPRLISCIII0507-24 | ERS27214726 | 2024 | MAN | EXT | | 162 | 801 | 15063 | 8 | 70 | 4 | 5 | 6 | 116 | 6 | 1_0_2 | . | . | . | . | . | | | |
| SPRLISCIII0535-19 | ERS27214727 | 2019 | WOMAN | CAT | | 162 | 801 | 15063 | 8 | 70 | 4 | 5 | 6 | 116 | 6 | 1_0_2 | . | . | . | . | . | | | |
| SPRLISCIII0545-24 | ERS27214728 | 2024 | MAN | MAD | | 27 | 899 | 247 | 16 | 13 | 4 | 5 | 6 | 10 | 14 | 0_0_3 | . | . | . | . | . | | | PI-1 |
| SPRLISCIII0572-22 | ERS27214729 | 2022 | MAN | AND | SEV | 162 | 801 | 15063 | 8 | 70 | 4 | 5 | 6 | 116 | 6 | 1_0_2 | . | . | . | . | . | | | |
| SPRLISCIII0580-09 | ERS27214730 | 2009 | WOMAN | CAT | | 70 | 259 | 259 | 24 | 5 | 35 | 1 | 15 | 12 | 43 | 2_44_2 | . | . | . | . | . | | PPH010 | PI-1 |
| SPRLISCIII0582-23DU | ERS27214731 | 2023 | MAN | AND | SEV | 162 | 801 | 15063 | 8 | 70 | 4 | 5 | 6 | 116 | 6 | 1_0_2 | . | . | . | . | . | | | |
| SPRLISCIII0587-18 | ERS27214732 | 2018 | MAN | MAD | | 182 | 2941 | 2941 | 97 | 5 | 1 | 1 | 36 | 1 | 14 | 2_4_2 | . | . | . | . | *tet*(M)_12 | Tn*916*-family | PPH010 | PI-1 |
| SPRLISCIII0649-19 | ERS27214733 | 2019 | MAN | CAT | | 27 | 244 | 244 | 16 | 2 | 4 | 1 | 6 | 10 | 18 | 0_0_3 | *cat*(pC194)_1 | *erm*(B)_18 | . | . | tet(M)_8 | Tn*916*-family | | PI-1 |
| SPRLISCIII0684-23 | ERS27214734 | 2023 | MAN | AND | SEV | 162 | 801 | 15063 | 8 | 70 | 4 | 5 | 6 | 116 | 6 | 1_0_2 | . | . | . | . | . | | | |
| SPRLISCIII0704-24 | ERS27214735 | 2024 | MAN | AND | SEV | 162 | 801 | 15063 | 8 | 70 | 4 | 5 | 6 | 116 | 6 | 1_0_2 | . | . | . | . | . | | | |
| SPRLISCIII0724-18 | ERS27214736 | 2018 | MAN | ARA | | 27 | 205 | 205 | 10 | 5 | 4 | 5 | 13 | 10 | 18 | 0_0_3 | . | . | . | . | . | | | PI-1 |
| SPRLISCIII0724-21 | ERS27214737 | 2021 | MAN | VAL | | 162 | 801 | 801 | 8 | 70 | 4 | 1 | 6 | 116 | 6 | 1_0_2 | . | . | *mef*(A)_2 | *msr*(D)_2 | . | ND | | |
| SPRLISCIII0726-24 | ERS27214738 | 2024 | MAN | AND | | 162 | 801 | 15063 | 8 | 70 | 4 | 5 | 6 | 116 | 6 | 1_0_2 | . | . | . | . | . | | | |
| SPRLISCIII0749-19 | ERS27214739 | 2019 | WOMAN | GAL | | 162 | 801 | 15063 | 8 | 70 | 4 | 5 | 6 | 116 | 6 | 1_0_2 | . | . | . | . | . | | | |
| SPRLISCIII0766-18 | ERS27214740 | 2018 | MAN | VAS | | 86 | 2712 | 2712 | 15 | 11 | 54 | 5 | 103 | 27 | 9 | 0_0_0 | . | . | . | . | . | | | |
| SPRLISCIII0770-09 | ERS27214741 | 2009 | NA | GAL | | 27 | 899 | 246 | 16 | 13 | 4 | 5 | 6 | 10 | 18 | 0_0_3 | . | . | . | . | . | | | PI-1 |
| SPRLISCIII0779-24 | ERS27214742 | 2024 | MAN | AND | SEV | 162 | 801 | 15063 | 8 | 70 | 4 | 5 | 6 | 116 | 6 | 1_0_2 | . | . | . | . | . | | | |
| SPRLISCIII0780-24 | ERS27214743 | 2024 | MAN | AND | SEV | 162 | 801 | 15063 | 8 | 70 | 4 | 5 | 6 | 116 | 6 | 1_0_2 | . | . | . | . | . | | | |
| SPRLISCIII0800-23 | ERS27214744 | 2023 | MAN | MUR | | 27 | 205 | 205 | 10 | 5 | 4 | 5 | 13 | 10 | 18 | 0_0_3 | . | . | . | . | . | | | PI-1 |
| SPRLISCIII0813-21 | ERS27214745 | 2021 | MAN | VAL | | 182 | 2941 | 9197 | 97 | 5 | 1 | 226 | 36 | 1 | 14 | 2_4_2 | . | . | . | . | . | | | PI-1 |

| | | | | | | | | | | | | | | | | | | | | | | | | |
|---|---|---|---|---|---|---|---|---|---|---|---|---|---|---|---|---|---|---|---|---|---|---|---|---|
| SPRLISCIII0831-22 | ERS27214746 | 2022 | MAN | CAT | | 162 | 801 | 13022 | 8 | 32 | 4 | 1 | 6 | 4 | 6 | NEW_0_2 | . | . | . | . | . | | | |
| SPRLISCIII0842-24 | ERS27214747 | 2024 | MAN | CAT | | 162 | 801 | 13022 | 8 | 32 | 4 | 1 | 6 | 4 | 6 | NEW_0_2 | . | . | . | . | . | | | |
| SPRLISCIII0847-23 | ERS27214748 | 2023 | MAN | CAT | | 162 | 801 | 13022 | 8 | 32 | 4 | 1 | 6 | 4 | 6 | NEW_0_2 | . | . | . | . | . | | | |
| SPRLISCIII0849-22 | ERS27214749 | 2022 | MAN | AND | SEV | 162 | 801 | 15063 | 8 | 70 | 4 | 5 | 6 | 116 | 6 | 1_0_2 | . | . | *mef*(A)_2 | *msr*(D)_2 | . | ND | ND | |
| SPRLISCIII0850-22 | ERS27214750 | 2022 | MAN | AND | SEV | 162 | 801 | 15063 | 8 | 70 | 4 | 5 | 6 | 116 | 6 | 1_0_2 | . | . | . | . | . | | | |
| SPRLISCIII0855-24V | ERS27214751 | 2024 | MAN | VAS | | 162 | 801 | 15063 | 8 | 70 | 4 | 5 | 6 | 116 | 6 | 1_0_2 | . | . | . | . | . | | | |
| SPRLISCIII0857-22 | ERS27214752 | 2022 | MAN | BAL | | 162 | 801 | 14136 | 8 | 12 | 4 | 1 | 17 | 116 | 72 | 23_0_2 | . | . | . | . | . | | | |
| SPRLISCIII0859-24V | ERS27214753 | 2024 | MAN | VAS | | 182 | 2941 | 9197 | 97 | 5 | 1 | 226 | 36 | 1 | 14 | 2_4_2 | . | . | . | . | . | | | PI-1 |
| SPRLISCIII0861-24V | ERS27214754 | 2024 | MAN | VAS | | 182 | 2941 | 9197 | 97 | 5 | 1 | 226 | 36 | 1 | 14 | 2_4_2 | . | . | . | . | . | | | PI-1 |
| SPRLISCIII0862-23 | ERS27214755 | 2023 | MAN | CAN | | 162 | 801 | 15063 | 8 | 70 | 4 | 5 | 6 | 116 | 6 | 1_0_2 | . | . | . | . | . | | | |
| SPRLISCIII0872-24 | ERS27214756 | 2024 | MAN | CAT | | 162 | 801 | 13022 | 8 | 32 | 4 | 1 | 6 | 4 | 6 | NEW_0_2 | . | . | . | . | . | | | |
| SPRLISCIII0876-09 | ERS27214757 | 2009 | MAN | GAL | | 70 | 1221 | NEW | 7 | 11 | 1 | 5 | 15 | 12 | 8 | 2_0_3 | . | . | . | . | . | | | PI-1 |
| SPRLISCIII0885-23 | ERS27214758 | 2023 | MAN | CAT | | 162 | 801 | 13022 | 8 | 32 | 4 | 1 | 6 | 4 | 6 | NEW_0_2 | . | . | . | . | . | | | |
| SPRLISCIII0899-21 | ERS27214759 | 2021 | MAN | BAL | | 162 | 801 | 14136 | 8 | 12 | 4 | 1 | 17 | 116 | 72 | 23_0_2 | . | . | . | . | . | | | |
| SPRLISCIII0900-22 | ERS27214760 | 2022 | MAN | AND | | 27 | 205 | 205 | 10 | 5 | 4 | 5 | 13 | 10 | 18 | 0_0_3 | . | . | . | . | . | | | PI-1 |
| SPRLISCIII0920-24 | ERS27214761 | 2024 | WOMAN | AND | | 162 | 801 | 15063 | 8 | 70 | 4 | 5 | 6 | 116 | 6 | 1_0_2 | . | . | . | . | . | | | |
| SPRLISCIII0922-21 | ERS27214762 | 2021 | MAN | VAL | | 162 | 801 | 801 | 8 | 70 | 4 | 1 | 6 | 116 | 6 | 1_0_2 | . | . | *mef*(A)_2 | *msr*(D)_2 | . | ND | | |
| SPRLISCIII0934-23 | ERS27214763 | 2023 | WOMAN | AND | | 162 | 801 | 15063 | 8 | 70 | 4 | 5 | 6 | 116 | 6 | 1_0_2 | . | . | . | . | . | | | |
| SPRLISCIII0934-24 | ERS27214764 | 2024 | MAN | CAT | | 162 | 801 | 13022 | 8 | 32 | 4 | 1 | 6 | 4 | 6 | NEW_0_2 | . | . | . | . | . | | | |
| SPRLISCIII0939-23 | ERS27214765 | 2023 | MAN | C-L | | NA | 1866 | 1866 | 1 | 11 | 1 | 15 | 103 | 1 | 70 | 0_0_0 | . | . | . | . | . | | | |
| SPRLISCIII0959-23 | ERS27214766 | 2023 | MAN | AND | | 162 | 801 | 15063 | 8 | 70 | 4 | 5 | 6 | 116 | 6 | 1_0_2 | . | . | . | . | . | | | |
| SPRLISCIII0963-24 | ERS27214767 | 2024 | MAN | AND | SEV | 162 | 801 | 15063 | 8 | 70 | 4 | 5 | 6 | 116 | 6 | 1_0_2 | . | . | . | . | . | | | |

| | | | | | | | | | | | | | | | | | | | | | | | | |
|---|---|---|---|---|---|---|---|---|---|---|---|---|---|---|---|---|---|---|---|---|---|---|---|---|
| SPRLISCIII0965-24 | ERS27214768 | 2024 | MAN | AND | SEV | 162 | 801 | 15063 | 8 | 70 | 4 | 5 | 6 | 116 | 6 | 1_0_2 | . | . | . | . | . | | | |
| SPRLISCIII0966-24V | ERS27214769 | 2024 | MAN | AND | SEV | 162 | 801 | 15063 | 8 | 70 | 4 | 5 | 6 | 116 | 6 | 1_0_2 | . | . | . | . | . | | | |
| SPRLISCIII0973-24B | ERS27214770 | 2024 | MAN | CAN | | 182 | 2941 | 9197 | 97 | 5 | 1 | 226 | 36 | 1 | 14 | 2_4_2 | . | . | . | . | . | | PPH010 | PI-1 |
| SPRLISCIII0974-21 | ERS27214771 | 2021 | MAN | AND | SEV | 162 | 801 | 15063 | 8 | 70 | 4 | 5 | 6 | 116 | 6 | 1_0_2 | . | . | . | . | . | | | |
| SPRLISCIII0975-22 | ERS27214772 | 2022 | WOMAN | C-L | | 162 | 801 | 1222 | 8 | 32 | 4 | 1 | 6 | 116 | 6 | 23_40_2 | . | . | . | . | . | | | |
| SPRLISCIII0980-23 | ERS27214773 | 2023 | MAN | AND | | 162 | 801 | 15063 | 8 | 70 | 4 | 5 | 6 | 116 | 6 | 1_0_2 | . | . | . | . | . | | | |
| SPRLISCIII0993-09 | ERS27214774 | 2009 | WOMAN | CAT | | 27 | 205 | 206 | 10 | 5 | 17 | 5 | 13 | 10 | 18 | 0_0_3 | . | . | . | . | . | | | PI-1 |
| SPRLISCIII1005-22 | ERS27214775 | 2022 | MAN | VAS | | 162 | 801 | 1222 | 8 | 32 | 4 | 1 | 6 | 116 | 6 | 23_40_2 | . | . | . | . | . | | | |
| SPRLISCIII1005-24 | ERS27214776 | 2024 | MAN | AND | SEV | 162 | 801 | 15063 | 8 | 70 | 4 | 5 | 6 | 116 | 6 | 1_0_2 | . | . | . | . | . | | | |
| SPRLISCIII1007-24 | ERS27214777 | 2024 | WOMAN | AND | SEV | 162 | 801 | 15063 | 8 | 70 | 4 | 5 | 6 | 116 | 6 | 1_0_2 | . | . | . | . | . | | | |
| SPRLISCIII1036-22 | ERS27214778 | 2022 | MAN | AND | SEV | 162 | 801 | 15063 | 8 | 70 | 4 | 5 | 6 | 116 | 6 | 1_0_2 | . | . | . | . | . | | | |
| SPRLISCIII1053-24V | ERS27214779 | 2024 | MAN | AND | SEV | 162 | 801 | 15063 | 8 | 70 | 4 | 5 | 6 | 116 | 6 | 1_0_2 | . | . | . | . | . | | | |
| SPRLISCIII1074-23 | ERS27214780 | 2023 | MAN | AND | | 162 | 801 | 15063 | 8 | 70 | 4 | 5 | 6 | 116 | 6 | 1_0_2 | . | . | . | . | . | | | |
| SPRLISCIII1115-24 | ERS27214781 | 2024 | WOMAN | AND | | 162 | 801 | 15063 | 8 | 70 | 4 | 5 | 6 | 116 | 6 | 1_0_2 | . | . | . | . | . | | | |
| SPRLISCIII1134-22 | ERS27214782 | 2022 | MAN | MUR | | NA | 1866 | NEW | 1 | 11 | 1 | 15 | ~103 | 1 | 70 | 0_0_0 | . | . | . | . | . | | | |
| SPRLISCIII1150-24 | ERS27214783 | 2024 | MAN | ARA | | 162 | 801 | 13022 | 8 | 32 | 4 | 1 | 6 | 4 | 6 | NEW_0_2 | . | . | . | . | . | | | |
| SPRLISCIII1157-24V | ERS27214784 | 2024 | MAN | AND | SEV | 162 | 801 | 15063 | 8 | 70 | 4 | 5 | 6 | 116 | 6 | 1_0_2 | . | . | . | . | . | | | |
| SPRLISCIII1170-23 | ERS27214785 | 2023 | MAN | VAS | | 182 | 2941 | 2941 | 97 | 5 | 1 | 1 | 36 | 1 | 14 | 2_4_2 | . | . | . | . | *tet*(M)_12 | Tn*916*-family | PPH010 | PI-1 |
| SPRLISCIII1174-22 | ERS27214786 | 2022 | MAN | AND | SEV | 162 | 801 | 15063 | 8 | 70 | 4 | 5 | 6 | 116 | 6 | 1_0_NEW | . | . | . | . | . | | | |
| SPRLISCIII1179-23 | ERS27214787 | 2023 | MAN | AND | SEV | 162 | 801 | 15063 | 8 | 70 | 4 | 5 | 6 | 116 | 6 | 1_0_2 | . | . | . | . | . | | | |
| SPRLISCIII1181-15 | ERS27214788 | 2015 | MAN | CAT | | 162 | 801 | 801 | 8 | 70 | 4 | 1 | 6 | 116 | 6 | 1_0_2 | . | . | . | . | . | | | |
| SPRLISCIII1183-24 | ERS27214789 | 2024 | MAN | AND | | 162 | 801 | 15063 | 8 | 70 | 4 | 5 | 6 | 116 | 6 | 1_0_2 | . | . | . | . | . | | | |

| | | | | | | | | | | | | | | | | | | | | | | | | |
|---|---|---|---|---|---|---|---|---|---|---|---|---|---|---|---|---|---|---|---|---|---|---|---|---|
| SPRLISCIII1195-24 | ERS27214790 | 2024 | MAN | AND | SEV | 162 | 801 | 15063 | 8 | 70 | 4 | 5 | 6 | 116 | 6 | 1_0_2 | . | . | . | . | . | | | |
| SPRLISCIII1244-09 | ERS27214791 | 2009 | MAN | MUR | | 27 | 205 | 205 | 10 | 5 | 4 | 5 | 13 | 10 | 18 | 0_0_3 | . | . | . | . | . | | | PI-1 |
| SPRLISCIII1244-23 | ERS27214792 | 2023 | WOMAN | CAT | | 162 | 801 | 13022 | 8 | 32 | 4 | 1 | 6 | 4 | 6 | NEW_0_2 | . | . | . | . | . | | | |
| SPRLISCIII1246-18 | ERS27214793 | 2018 | MAN | AND | SEV | NA | 1866 | 1866 | 1 | 11 | 1 | 15 | 103 | 1 | 70 | 0_0_0 | . | . | . | . | . | | Streptococcus phage Javan340-like | |
| SPRLISCIII1249-24 | ERS27214794 | 2024 | MAN | C-L | | 162 | 801 | 1222 | 8 | 32 | 4 | 1 | 6 | 116 | 6 | 23_40_2 | . | . | . | . | . | | | |
| SPRLISCIII1269-19 | ERS27214795 | 2019 | MAN | CAT | | 27 | 205 | 205 | 10 | 5 | 4 | 5 | 13 | 10 | 18 | 0_0_3 | . | . | . | . | . | | | PI-1 |
| SPRLISCIII1271-23 | ERS27214796 | 2023 | MAN | CAT | | 27 | 205 | 205 | 10 | 5 | 4 | 5 | 13 | 10 | 18 | NEW_0_3 | . | . | . | . | . | | | PI-1 |
| SPRLISCIII1275-24 | ERS27214797 | 2024 | MAN | C-M | | 162 | 801 | 15063 | 8 | 70 | 4 | 5 | 6 | 116 | 6 | 1_0_2 | . | . | . | . | . | | | |
| SPRLISCIII1284-09 | ERS27214798 | 2009 | WOMAN | BAL | | 27 | 899 | 247 | 16 | 13 | 4 | 5 | 6 | 10 | 14 | 0_0_3 | . | . | . | . | . | | | PI-1 |
| SPRLISCIII1291-24 | ERS27214799 | 2024 | MAN | AND | | 162 | 801 | 15063 | 8 | 70 | 4 | 5 | 6 | 116 | 6 | 1_0_2 | . | . | . | . | . | | | |
| SPRLISCIII1296-24 | ERS27214800 | 2024 | MAN | CAT | | 162 | 19102 | 19102 | 8 | 5 | 9 | 1 | 6 | 116 | 6 | 23_0_2 | . | . | . | . | . | | | |
| SPRLISCIII1315-22 | ERS27214801 | 2022 | WOMAN | MAD | | 27 | 899 | 247 | 16 | 13 | 4 | 5 | 6 | 10 | 14 | 0_0_3 | . | . | . | . | . | | | PI-1 |
| SPRLISCIII1324-19 | ERS27214802 | 2019 | MAN | GAL | | 162 | 801 | 15063 | 8 | 70 | 4 | 5 | 6 | 116 | 6 | 1_0_2 | . | . | . | . | . | | | |
| SPRLISCIII1348-22 | ERS27214803 | 2022 | WOMAN | BAL | | NA | 1866 | 1866 | 1 | 11 | 1 | 15 | 103 | 1 | 70 | 0_0_0 | . | . | . | . | . | | | |
| SPRLISCIII1351-23 | ERS27214804 | 2023 | MAN | MUR | | 162 | 801 | 801 | 8 | 70 | 4 | 1 | 6 | 116 | 6 | 1_0_2 | . | . | *mef*(A)_2 | *msr*(D)_2 | . | ND | | |
| SPRLISCIII1360-22 | ERS27214805 | 2022 | MAN | CAB | | 162 | 801 | 1222 | 8 | 32 | 4 | 1 | 6 | 116 | 6 | 23_40_2 | . | . | . | . | . | | | |
| SPRLISCIII1361-24 | ERS27214806 | 2024 | WOMAN | CAT | | 162 | 5872 | 5872 | 2 | 128 | 4 | 1 | 14 | 1 | 72 | 0_0_2 | . | *erm*(B)_18 | . | . | *tet*(M)_5 | Tn*1549*-family | | |
| SPRLISCIII1372-24 | ERS27214807 | 2024 | MAN | CAT | | 162 | 801 | 13022 | 8 | 32 | 4 | 1 | 6 | 4 | 6 | NEW_0_2 | . | . | . | . | . | | | |
| SPRLISCIII1374-18 | ERS27214808 | 2018 | MAN | BAL | | 162 | 801 | NEW | 1 | 8 | 4 | 1 | 6 | 116 | ~31 | 23_4_2 | . | . | . | . | . | | | |
| SPRLISCIII1382-24 | ERS27214809 | 2024 | MAN | AND | SEV | 162 | 801 | 15063 | 8 | 70 | 4 | 5 | 6 | 116 | 6 | 1_0_2 | . | . | . | . | . | | | |
| SPRLISCIII1383-24 | ERS27214810 | 2024 | WOMAN | AND | | 162 | 801 | 15063 | 8 | 70 | 4 | 5 | 6 | 116 | 6 | 1_0_2 | . | . | . | . | . | | | |

| | | | | | | | | | | | | | | | | | | | | | | | |
|---|---|---|---|---|---|---|---|---|---|---|---|---|---|---|---|---|---|---|---|---|---|---|---|
| SPRLISCIII1400-23 | ERS27214811 | 2023 | MAN | AND | SEV | 162 | 801 | 15063 | 8 | 70 | 4 | 5 | 6 | 116 | 6 | 1_0_2 | . | . | . | . | . | | |
| SPRLISCIII1426-23 | ERS27214812 | 2023 | MAN | AND | | 162 | 801 | 15063 | 8 | 70 | 4 | 5 | 6 | 116 | 6 | 1_0_2 | . | . | . | . | . | | |
| SPRLISCIII1434-09 | ERS27214813 | 2009 | MAN | C-L | | 162 | 801 | 1222 | 8 | 32 | 4 | 1 | 6 | 116 | 6 | 23_40_2 | . | . | . | . | . | | |
| SPRLISCIII1440-23 | ERS27214814 | 2023 | MAN | AND | | 162 | 801 | 15063 | 8 | 70 | 4 | 5 | 6 | 116 | 6 | 1_0_2 | . | . | . | . | . | | |
| SPRLISCIII1456-23 | ERS27214815 | 2023 | MAN | BAL | | 162 | 801 | 1222 | 8 | 32 | 4 | 1 | 6 | 116 | 6 | NEW_0_2 | . | . | . | . | . | | |
| SPRLISCIII1459-24 | ERS27214816 | 2024 | MAN | AND | | 162 | 801 | 15063 | 8 | 70 | 4 | 5 | 6 | 116 | 6 | 1_0_2 | . | . | . | . | . | | |
| SPRLISCIII1460-24 | ERS27214817 | 2024 | WOMAN | CAT | | 162 | 801 | 15063 | 8 | 70 | 4 | 5 | 6 | 116 | 6 | 1_0_2 | . | . | . | . | . | | |
| SPRLISCIII1465-09 | ERS27214818 | 2009 | MAN | VAL | | 162 | 801 | NEW | 8 | 8 | 4 | 1 | 6 | 116 | ~6 | 23_0_2 | *catQ*_1 | . | *mef*(A)_1 | *msr*(D)_2 | *tet*(M)_2 | ICE*Spn*_1465 | |
| SPRLISCIII1471-15 | ERS27214819 | 2015 | MAN | CAT | | 736 | 6864 | 6864 | 118 | 60 | 4 | 15 | 25 | 14 | 9 | 150_350_77 | . | . | . | . | . | | |
| SPRLISCIII1472-18 | ERS27214820 | 2018 | MAN | CAT | | 86 | 2712 | 2712 | 15 | 11 | 54 | 5 | 103 | 27 | 9 | 0_0_0 | . | . | . | . | . | | |
| SPRLISCIII1474-23 | ERS27214821 | 2023 | MAN | AND | | 162 | 801 | 15063 | 8 | 70 | 4 | 5 | 6 | 116 | 6 | 1_0_2 | . | . | . | . | . | | |
| SPRLISCIII1479-23 | ERS27214822 | 2023 | MAN | AND | SEV | 162 | 801 | 15063 | 8 | 70 | 4 | 5 | 6 | 116 | 6 | 1_0_2 | . | . | . | . | . | | |
| SPRLISCIII1508-23 | ERS27214823 | 2023 | MAN | CAT | | 162 | 801 | 13022 | 8 | 32 | 4 | 1 | 6 | 4 | 6 | NEW_0_2 | . | . | . | . | . | | |
| SPRLISCIII1515-22 | ERS27214824 | 2022 | WOMAN | CAT | | 162 | 801 | 13022 | 8 | 32 | 4 | 1 | 6 | 4 | 6 | NEW_0_2 | . | . | . | . | . | | |
| SPRLISCIII1519-22 | ERS27214825 | 2022 | MAN | MAD | | 27 | 205 | 205 | 10 | 5 | 4 | 5 | 13 | 10 | 18 | 0_0_3 | . | . | . | . | . | | PI-1 |
| SPRLISCIII1530-22 | ERS27214826 | 2022 | MAN | MAD | | 162 | 801 | 15063 | 8 | 70 | 4 | 5 | 6 | 116 | 6 | 1_0_2 | . | . | . | . | . | | |
| SPRLISCIII1547-19 | ERS27214827 | 2019 | WOMAN | CAT | | 27 | 205 | 205 | 10 | 5 | 4 | 5 | 13 | 10 | 18 | 0_0_3 | . | . | . | . | . | | PI-1 |
| SPRLISCIII1562-23 | ERS27214828 | 2023 | MAN | C-M | | 162 | 801 | 15063 | 8 | 70 | 4 | 5 | 6 | 116 | 6 | 1_0_2 | . | . | . | . | . | | |
| SPRLISCIII1569-24 | ERS27214829 | 2024 | MAN | AND | | 162 | 801 | 15063 | 8 | 70 | 4 | 5 | 6 | 116 | 6 | 1_0_2 | . | . | . | . | . | | |
| SPRLISCIII1570-15 | ERS27214830 | 2015 | MAN | CAT | | 162 | 801 | 13022 | 8 | 32 | 4 | 1 | 6 | 4 | 6 | NEW_0_2 | . | . | . | . | . | | |
| SPRLISCIII1592-22 | ERS27214831 | 2022 | MAN | AND | SEV | 162 | 801 | 15063 | 8 | 70 | 4 | 5 | 6 | 116 | 6 | 1_0_2 | . | . | . | . | . | | |
| SPRLISCIII1606-23 | ERS27214832 | 2023 | WOMAN | BAL | | NA | 1866 | 1866 | 1 | 11 | 1 | 15 | 103 | 1 | 70 | 0_0_0 | . | . | . | . | . | | |

| | | | | | | | | | | | | | | | | | | | | | | | |
|---|---|---|---|---|---|---|---|---|---|---|---|---|---|---|---|---|---|---|---|---|---|---|---|
| SPRLISCIII1607-19 | ERS27214833 | 2019 | MAN | CAT | | 162 | 801 | 13022 | 8 | 32 | 4 | 1 | 6 | 4 | 6 | NEW_0_2 | . | . | . | . | . | | |
| SPRLISCIII1616-19 | ERS27214834 | 2019 | MAN | VAS | | 162 | 801 | 1222 | 8 | 32 | 4 | 1 | 6 | 116 | 6 | 23_40_2 | . | . | . | . | . | | |
| SPRLISCIII1635-24 | ERS27214835 | 2024 | MAN | AND | SEV | 162 | 801 | 15063 | 8 | 70 | 4 | 5 | 6 | 116 | 6 | 1_0_2 | . | . | . | . | . | | |
| SPRLISCIII1636-24 | ERS27214836 | 2024 | MAN | AND | SEV | 162 | 801 | 15063 | 8 | 70 | 4 | 5 | 6 | 116 | 6 | 1_0_2 | . | . | . | . | . | | |
| SPRLISCIII1639-24 | ERS27214837 | 2024 | MAN | AND | SEV | 162 | 801 | 15063 | 8 | 70 | 4 | 5 | 6 | 116 | 6 | 1_0_2 | . | . | . | . | . | | |
| SPRLISCIII1654-09 | ERS27214838 | 2009 | WOMAN | GAL | | 162 | 801 | 1222 | 8 | 32 | 4 | 1 | 6 | 116 | 6 | 23_40_2 | . | . | . | . | . | | |
| SPRLISCIII1683-24 | ERS27214839 | 2024 | WOMAN | MAD | | 27 | 899 | 247 | 16 | 13 | 4 | 5 | 6 | 10 | 14 | 0_0_3 | . | . | . | . | . | | PI-1 |
| SPRLISCIII1711-23 | ERS27214840 | 2023 | MAN | AND | SEV | 162 | 801 | 15063 | 8 | 70 | 4 | 5 | 6 | 116 | 6 | 1_0_2 | . | . | . | . | . | | |
| SPRLISCIII1728-23 | ERS27214841 | 2023 | WOMAN | AND | SEV | 162 | 801 | 15063 | 8 | 70 | 4 | 5 | 6 | 116 | 6 | 1_0_2 | . | . | . | . | . | | |
| SPRLISCIII1733-24 | ERS27214842 | 2024 | MAN | AST | | 162 | 801 | 801 | 8 | 70 | 4 | 1 | 6 | 116 | 6 | 1_0_2 | . | . | . | . | . | | |
| SPRLISCIII1734-24 | ERS27214843 | 2024 | MAN | AND | | 162 | 801 | 15063 | 8 | 70 | 4 | 5 | 6 | 116 | 6 | 1_0_2 | . | . | . | . | . | | |
| SPRLISCIII1743-24 | ERS27214844 | 2024 | WOMAN | MUR | | 162 | 801 | 15063 | 8 | 70 | 4 | 5 | 6 | 116 | 6 | 1_0_2 | . | . | . | . | . | | |
| SPRLISCIII1754-18 | ERS27214845 | 2018 | MAN | C-L | | 162 | 801 | 15063 | 8 | 70 | 4 | 5 | 6 | 116 | 6 | 1_0_2 | . | . | . | . | . | | |
| SPRLISCIII1761-24 | ERS27214846 | 2024 | MAN | AND | | 162 | 801 | 15063 | 8 | 70 | 4 | 5 | 6 | 116 | 6 | 1_0_2 | . | . | . | . | . | | |
| SPRLISCIII1772-24 | ERS27214847 | 2024 | WOMAN | AND | | 162 | 801 | 15063 | 8 | 70 | 4 | 5 | 6 | 116 | 6 | 1_0_2 | . | . | . | . | . | | |
| SPRLISCIII1811-24 | ERS27214848 | 2024 | WOMAN | AND | SEV | 162 | 801 | 15063 | 8 | 70 | 4 | 5 | 6 | 116 | 6 | 1_0_2 | . | . | . | . | . | | |
| SPRLISCIII1819-24 | ERS27214849 | 2024 | MAN | AND | SEV | 162 | 801 | 15063 | 8 | 70 | 4 | 5 | 6 | 116 | 6 | 1_0_12 | . | . | . | . | . | | |
| SPRLISCIII1820-24 | ERS27214850 | 2024 | WOMAN | AND | SEV | 162 | 801 | 15063 | 8 | 70 | 4 | 5 | 6 | 116 | 6 | 1_0_2 | . | . | . | . | . | | |
| SPRLISCIII1821-24 | ERS27214851 | 2024 | WOMAN | AND | SEV | 162 | 801 | 15063 | 8 | 70 | 4 | 5 | 6 | 116 | 6 | 1_0_2 | . | . | . | . | . | | |
| SPRLISCIII1822-24 | ERS27214852 | 2024 | MAN | AND | SEV | 162 | 801 | 15063 | 8 | 70 | 4 | 5 | 6 | 116 | 6 | 1_0_2 | . | . | . | . | . | | |
| SPRLISCIII1830-24 | ERS27214853 | 2024 | MAN | MAD | | 162 | 801 | 801 | 8 | 70 | 4 | 1 | 6 | 116 | 6 | 1_0_2 | . | . | . | . | . | | |
| SPRLISCIII1872-24 | ERS27214854 | 2024 | MAN | AND | SEV | 162 | 801 | 801 | 8 | 70 | 4 | 1 | 6 | 116 | 6 | 1_0_2 | . | . | *mef*(A)_2 | *msr*(D)_2 | . | ND | |

| | | | | | | | | | | | | | | | | | | | | | | | | |
|---|---|---|---|---|---|---|---|---|---|---|---|---|---|---|---|---|---|---|---|---|---|---|---|---|
| SPRLISCIII1893-24 | ERS27214855 | 2024 | WOMAN | MUR | | 162 | 801 | 15063 | 8 | 70 | 4 | 5 | 6 | 116 | 6 | 1_0_2 | . | . | . | . | . | | | |
| SPRLISCIII1894-24 | ERS27214856 | 2024 | MAN | EXT | | 162 | 801 | 15063 | 8 | 70 | 4 | 5 | 6 | 116 | 6 | 1_0_2 | . | . | . | . | . | | PPH010 | |
| SPRLISCIII1896-19 | ERS27214857 | 2019 | WOMAN | CAT | | 162 | 801 | 13022 | 8 | 32 | 4 | 1 | 6 | 4 | 6 | NEW_0_2 | . | . | . | . | . | | | |
| SPRLISCIII1914-22 | ERS27214858 | 2022 | MAN | CAN | | 162 | 801 | 15063 | 8 | 70 | 4 | 5 | 6 | 116 | 6 | 1_0_2 | . | . | . | . | . | | | |
| SPRLISCIII1916-24 | ERS27214859 | 2024 | MAN | AND | SEV | 162 | 801 | 15063 | 8 | 70 | 4 | 5 | 6 | 116 | 6 | 1_0_2 | . | . | . | . | . | | | |
| SPRLISCIII1922-23 | ERS27214860 | 2023 | WOMAN | CAT | | 427 | 10172 | 10172 | 1 | 8 | 9 | 15 | 6 | 58 | 70 | 3_0_0 | . | . | . | . | . | | | |
| SPRLISCIII1927-22 | ERS27214861 | 2022 | MAN | AND | SEV | 162 | 801 | 15063 | 8 | 70 | 4 | 5 | 6 | 116 | 6 | 1_0_2 | . | . | . | . | . | | | |
| SPRLISCIII1932-23 | ERS27214862 | 2023 | MAN | CAT | | 162 | 801 | 15063 | 8 | 70 | 4 | 5 | 6 | 116 | 6 | 1_0_2 | . | . | . | . | . | | | |
| SPRLISCIII1966-23 | ERS27214863 | 2023 | MAN | AND | SEV | 162 | 801 | 15063 | 8 | 70 | 4 | 5 | 6 | 116 | 6 | 1_0_2 | . | . | . | . | . | | | |
| SPRLISCIII1967-18 | ERS27214864 | 2018 | MAN | CAT | | 162 | 801 | 13022 | 8 | 32 | 4 | 1 | 6 | 4 | 6 | NEW_0_2 | . | . | . | . | . | | | |
| SPRLISCIII1972-23 | ERS27214865 | 2023 | MAN | AND | SEV | 162 | 801 | 15063 | 8 | 70 | 4 | 5 | 6 | 116 | 6 | 1_0_2 | . | . | . | . | . | | | |
| SPRLISCIII1977-23 | ERS27214866 | 2023 | MAN | AND | SEV | 162 | 801 | 15063 | 8 | 70 | 4 | 5 | 6 | 116 | 6 | 1_0_2 | . | . | . | . | . | | | |
| SPRLISCIII1989-24 | ERS27214867 | 2024 | MAN | C-L | | 27 | 899 | 247 | 16 | 13 | 4 | 5 | 6 | 10 | 14 | 0_0_3 | . | . | . | . | . | | | PI-1 |
| SPRLISCIII1991-23 | ERS27214868 | 2023 | WOMAN | MAD | | 162 | 801 | 15063 | 8 | 70 | 4 | 5 | 6 | 116 | 6 | 1_0_2 | . | . | . | . | . | | | |
| SPRLISCIII1992-22 | ERS27214869 | 2022 | MAN | AND | SEV | 162 | 801 | 15063 | 8 | 70 | 4 | 5 | 6 | 116 | 6 | 1_0_2 | . | . | . | . | . | | | |
| SPRLISCIII1994-22 | ERS27214870 | 2022 | MAN | AND | SEV | 162 | 801 | 15063 | 8 | 70 | 4 | 5 | 6 | 116 | 6 | 1_0_2 | . | . | . | . | . | | | |
| SPRLISCIII1995-22 | ERS27214871 | 2022 | MAN | AND | SEV | 162 | 801 | 15063 | 8 | 70 | 4 | 5 | 6 | 116 | 6 | 1_0_2 | . | . | . | . | . | | | |
| SPRLISCIII1999-24 | ERS27214872 | 2024 | MAN | CAT | | 162 | 5872 | 5872 | 2 | 128 | 4 | 1 | 14 | 1 | 72 | 0_0_2 | . | *erm*(B)_18 | . | . | *tet*(M)_5 | Tn*1549*-family | | |
| SPRLISCIII2021-24 | ERS27214873 | 2024 | MAN | CAT | | 162 | 801 | 13022 | 8 | 32 | 4 | 1 | 6 | 4 | 6 | NEW_0_2 | . | . | . | . | . | | | |
| SPRLISCIII2028-23 | ERS27214874 | 2023 | MAN | AND | SEV | 162 | 801 | 15063 | 8 | 70 | 4 | 5 | 6 | 116 | 6 | 1_0_2 | . | . | . | . | . | | | |
| SPRLISCIII2032-22 | ERS27214875 | 2022 | MAN | CAT | | 162 | 801 | 13022 | 8 | 32 | 4 | 1 | 6 | 4 | 6 | NEW_0_2 | . | . | . | . | . | | | |
| SPRLISCIII2036-24 | ERS27214876 | 2024 | MAN | CAT | | 182 | 1729 | 1729 | 1 | 5 | 1 | 1 | 15 | 169 | 14 | 2_4_2 | . | . | . | . | . | | | |

| | | | | | | | | | | | | | | | | | | | | | | | |
|---|---|---|---|---|---|---|---|---|---|---|---|---|---|---|---|---|---|---|---|---|---|---|---|
| SPRLISCIII2039-22 | ERS27214877 | 2022 | MAN | AND | SEV | 162 | 801 | 15063 | 8 | 70 | 4 | 5 | 6 | 116 | 6 | 1_0_2 | . | . | . | . | . | | |
| SPRLISCIII2040-24 | ERS27214878 | 2024 | WOMAN | CAT | | 27 | 899 | 247 | 16 | 13 | 4 | 5 | 6 | 10 | 14 | 0_0_3 | . | . | . | . | . | | PI-1 |
| SPRLISCIII2047-23 | ERS27214879 | 2023 | MAN | VAS | | 528 | 6352 | 6352 | 15 | 11 | 146 | 15 | 11 | 3 | 9 | 0_0_0 | . | . | . | . | . | | |
| SPRLISCIII2065-22 | ERS27214880 | 2022 | MAN | C-L | | 162 | 801 | 1222 | 8 | 32 | 4 | 1 | 6 | 116 | 6 | 23_40_2 | . | . | . | . | . | | |
| SPRLISCIII2066-24 | ERS27214881 | 2024 | MAN | AND | SEV | 162 | 801 | 15063 | 8 | 70 | 4 | 5 | 6 | 116 | 6 | 1_0_2 | . | . | . | . | . | | |
| SPRLISCIII2067-24 | ERS27214882 | 2024 | WOMAN | AND | SEV | 162 | 801 | 15063 | 8 | 70 | 4 | 5 | 6 | 116 | 6 | 1_0_2 | . | . | . | . | . | | |
| SPRLISCIII2080-23 | ERS27214883 | 2023 | MAN | AND | SEV | 162 | 801 | 15063 | 8 | 70 | 4 | 5 | 6 | 116 | 6 | 1_0_2 | . | . | . | . | . | | |
| SPRLISCIII2084-24 | ERS27214884 | 2024 | MAN | C-L | | 162 | NA | NEW | 244 | 9 | 4 | 1 | 6 | 116 | ~18 | NEW_0_2 | . | . | . | . | *tet*(M)_1 | ND | |
| SPRLISCIII2093-18 | ERS27214885 | 2018 | MAN | VAL | | NA | 1866 | 1866 | 1 | 11 | 1 | 15 | 103 | 1 | 70 | 0_0_0 | . | . | . | . | . | | |
| SPRLISCIII2096-24 | ERS27214886 | 2024 | MAN | AND | SEV | 162 | 801 | 15063 | 8 | 70 | 4 | 5 | 6 | 116 | 6 | 1_0_2 | . | . | . | . | . | | |
| SPRLISCIII2098-23 | ERS27214887 | 2023 | WOMAN | VAS | | 528 | 6352 | 6352 | 15 | 11 | 146 | 15 | 11 | 3 | 9 | 0_0_0 | . | . | . | . | . | | |
| SPRLISCIII2106-23 | ERS27214888 | 2023 | MAN | AND | | 162 | 801 | 15063 | 8 | 70 | 4 | 5 | 6 | 116 | 6 | 1_0_2 | . | . | . | . | . | | |
| SPRLISCIII2117-19 | ERS27214889 | 2019 | MAN | AND | | 162 | 801 | 801 | 8 | 70 | 4 | 1 | 6 | 116 | 6 | 1_0_2 | . | . | . | . | . | | |
| SPRLISCIII2117-24 | ERS27214890 | 2024 | MAN | CAT | | 162 | 801 | 801 | 8 | 70 | 4 | 1 | 6 | 116 | 6 | 1_0_2 | . | . | . | . | . | | |
| SPRLISCIII2135-24 | ERS27214891 | 2024 | MAN | CAT | | 162 | 801 | 13022 | 8 | 32 | 4 | 1 | 6 | 4 | 6 | NEW_0_2 | . | . | . | . | . | | |
| SPRLISCIII2163-19 | ERS27214892 | 2019 | WOMAN | VAS | | 162 | 801 | 1222 | 8 | 32 | 4 | 1 | 6 | 116 | 6 | 23_40_2 | . | . | . | . | . | | |
| SPRLISCIII2184-09 | ERS27214893 | 2009 | MAN | CAT | | 70 | 1221 | 4795 | 7 | 5 | 1 | 5 | 15 | 12 | 8 | 2_0_3 | . | . | . | . | . | | PI-1 |
| SPRLISCIII2188-23 | ERS27214894 | 2023 | MAN | AND | SEV | 162 | 801 | 15063 | 8 | 70 | 4 | 5 | 6 | 116 | 6 | 1_0_2 | . | . | . | . | . | | |
| SPRLISCIII2196-18 | ERS27214895 | 2018 | MAN | CAT | | 70 | 1221 | 4795 | 7 | 5 | 1 | 5 | 15 | 12 | 8 | 2_0_3 | . | . | . | . | . | | PI-1 |
| SPRLISCIII2205-23 | ERS27214896 | 2023 | MAN | MAD | | 162 | 801 | 15063 | 8 | 70 | 4 | 5 | 6 | 116 | 6 | 1_0_2 | . | . | . | . | . | | |
| SPRLISCIII2206-22 | ERS27214897 | 2022 | WOMAN | ARA | | 182 | 2941 | 9197 | 97 | 5 | 1 | 226 | 36 | 1 | 14 | 2_4_2 | . | . | . | . | . | | PI-1 |
| SPRLISCIII2216-18 | ERS27214898 | 2018 | MAN | VAS | | 162 | 801 | 1222 | 8 | 32 | 4 | 1 | 6 | 116 | 6 | 23_40_2 | . | . | . | . | . | | |

| | | | | | | | | | | | | | | | | | | | | | | | | |
|---|---|---|---|---|---|---|---|---|---|---|---|---|---|---|---|---|---|---|---|---|---|---|---|---|
| SPRLISCIII2230-09 | ERS27214899 | 2009 | MAN | CAT | | 27 | 205 | 205 | 10 | 5 | 4 | 5 | 13 | 10 | 18 | 0_0_3 | . | . | . | . | . | | | PI-1 |
| SPRLISCIII2230-18 | ERS27214900 | 2018 | WOMAN | VAL | | NA | 1866 | 1866 | 1 | 11 | 1 | 15 | 103 | 1 | 70 | 0_0_0 | . | . | . | . | . | | | |
| SPRLISCIII2232-24 | ERS27214901 | 2024 | MAN | AND | | 162 | 801 | 15063 | 8 | 70 | 4 | 5 | 6 | 116 | 6 | 1_0_2 | . | . | . | . | . | | | |
| SPRLISCIII2239-24 | ERS27214902 | 2024 | WOMAN | AND | | 162 | 801 | 15063 | 8 | 70 | 4 | 5 | 6 | 116 | 6 | 1_0_2 | . | . | . | . | . | | | |
| SPRLISCIII2259-24 | ERS27214903 | 2024 | WOMAN | CAT | | 162 | 801 | 13022 | 8 | 32 | 4 | 1 | 6 | 4 | 6 | NEW_0_2 | . | . | . | . | . | | | |
| SPRLISCIII2261-22 | ERS27214904 | 2022 | MAN | AND | SEV | 162 | 801 | 15063 | 8 | 70 | 4 | 5 | 6 | 116 | 6 | 1_0_2 | . | . | . | . | . | | | |
| SPRLISCIII2264-22 | ERS27214905 | 2022 | WOMAN | AND | SEV | 162 | 801 | 15063 | 8 | 70 | 4 | 5 | 6 | 116 | 6 | 1_0_2 | . | . | . | . | . | | | |
| SPRLISCIII2267-22 | ERS27214906 | 2022 | MAN | AND | SEV | 162 | 801 | 15063 | 8 | 70 | 4 | 5 | 6 | 116 | 6 | 1_0_2 | . | . | . | . | . | | | |
| SPRLISCIII2278-24V | ERS27214907 | 2024 | WOMAN | CAT | | 331 | 19496 | 19496 | 7 | 88 | 103 | 6 | 36 | 142 | 161 | 12_2_418 | . | . | . | . | *tet*(M)_12 | Tn*916*-family | PPH085 | |
| SPRLISCIII2288-23 | ERS27214908 | 2023 | MAN | CAT | | 162 | 801 | 801 | 8 | 70 | 4 | 1 | 6 | 116 | 6 | 1_0_2 | . | . | . | . | . | | | |
| SPRLISCIII2299-24 | ERS27214909 | 2024 | WOMAN | EXT | | 182 | 2941 | 9197 | 97 | 5 | 1 | 226 | 36 | 1 | 14 | 2_4_2 | . | . | . | . | . | | | PI-1 |
| SPRLISCIII2313-22 | ERS27214910 | 2022 | MAN | AND | | 162 | 801 | 15063 | 8 | 70 | 4 | 5 | 6 | 116 | 6 | 1_0_2 | . | . | . | . | . | | | |
| SPRLISCIII2320-24 | ERS27214911 | 2024 | MAN | AND | | 162 | 801 | 801 | 8 | 70 | 4 | 1 | 6 | 116 | 6 | 1_0_2 | . | . | *mef*(A)_2 | *msr*(D)_2 | . | ND | | |
| SPRLISCIII2345-09 | ERS27214912 | 2009 | MAN | MAD | | 27 | 899 | 247 | 16 | 13 | 4 | 5 | 6 | 10 | 14 | 0_0_3 | . | . | . | . | . | | | PI-1 |
| SPRLISCIII2351-24 | ERS27214913 | 2024 | WOMAN | AND | SEV | 162 | 801 | 15063 | 8 | 70 | 4 | 5 | 6 | 116 | 6 | 1_0_2 | . | . | . | . | . | | | |
| SPRLISCIII2363-23 | ERS27214914 | 2023 | WOMAN | ARA | | 27 | 205 | 205 | 10 | 5 | 4 | 5 | 13 | 10 | 18 | 0_0_3 | . | . | . | . | . | | | PI-1 |
| SPRLISCIII2387-24V | ERS27214915 | 2024 | MAN | CAT | | 70 | 1221 | 1221 | 7 | 5 | 1 | 5 | 15 | 12 | 14 | 2_4_2 | . | . | . | . | . | | | PI-1 |
| SPRLISCIII2398-24 | ERS27214916 | 2024 | MAN | C-L | | 162 | 801 | 15063 | 8 | 70 | 4 | 5 | 6 | 116 | 6 | 1_0_2 | . | . | . | . | . | | | |
| SPRLISCIII2401-18 | ERS27214917 | 2018 | MAN | AND | | 331 | 1469 | 15410 | 8 | 88 | 103 | 18 | 9 | 142 | 161 | 12_2_418 | . | . | . | . | *tet*(M)_13 | Tn*916*-family | PPH080 | |
| SPRLISCIII2408-23 | ERS27214918 | 2023 | MAN | AND | | 162 | 801 | 15063 | 8 | 70 | 4 | 5 | 6 | 116 | 6 | 1_0_2 | . | . | . | . | . | | | |
| SPRLISCIII2420-24 | ERS27214919 | 2024 | MAN | NAV | | 27 | 899 | 247 | 16 | 13 | 4 | 5 | 6 | 10 | 14 | 0_0_3 | . | . | . | . | . | | | PI-1 |
| SPRLISCIII2439-24 | ERS27214920 | 2024 | MAN | AND | | 162 | 801 | 15063 | 8 | 70 | 4 | 5 | 6 | 116 | 6 | 1_0_2 | . | . | . | . | . | | | |

| | | | | | | | | | | | | | | | | | | | | | | | |
|---|---|---|---|---|---|---|---|---|---|---|---|---|---|---|---|---|---|---|---|---|---|---|---|
| SPRLISCIII2440-09 | ERS27214921 | 2009 | MAN | VAS | | 86 | 2036 | 2036 | 2 | 5 | 54 | 5 | 103 | 1 | 9 | 0_0_0 | . | . | . | . | . | PPH010; PPH100 | |
| SPRLISCIII2441-19 | ERS27214922 | 2019 | MAN | CAT | | 27 | 205 | 205 | 10 | 5 | 4 | 5 | 13 | 10 | 18 | 0_0_3 | . | . | . | . | . | | PI-1 |
| SPRLISCIII2441-24V | ERS27214923 | 2024 | MAN | ARA | | 427 | 10172 | 10172 | 1 | 8 | 9 | 15 | 6 | 58 | 70 | 3_0_0 | . | . | . | . | . | | |
| SPRLISCIII2447-23 | ERS27214924 | 2023 | MAN | AND | SEV | 162 | 801 | 15063 | 8 | 70 | 4 | 5 | 6 | 116 | 6 | 1_0_2 | . | . | . | . | . | | |
| SPRLISCIII2449-23 | ERS27214925 | 2023 | MAN | AND | | 162 | 801 | 15063 | 8 | 70 | 4 | 5 | 6 | 116 | 6 | 1_0_2 | . | . | . | . | . | | |
| SPRLISCIII2453-24 | ERS27214926 | 2024 | WOMAN | AND | SEV | 162 | 801 | 15063 | 8 | 70 | 4 | 5 | 6 | 116 | 6 | 1_0_2 | . | . | . | . | . | | |
| SPRLISCIII2455-09 | ERS27214927 | 2009 | MAN | VAL | | 27 | 205 | 205 | 10 | 5 | 4 | 5 | 13 | 10 | 18 | 0_0_3 | . | . | . | . | . | | PI-1 |
| SPRLISCIII2458-23 | ERS27214928 | 2023 | MAN | EXT | | 162 | 801 | 15063 | 8 | 70 | 4 | 5 | 6 | 116 | 6 | 1_0_2 | . | . | . | . | . | | |
| SPRLISCIII2459-24 | ERS27214929 | 2024 | MAN | AND | SEV | 162 | 801 | 15063 | 8 | 70 | 4 | 5 | 6 | 116 | 6 | 1_0_2 | . | . | . | . | . | | |
| SPRLISCIII2463-23 | ERS27214930 | 2023 | WOMAN | C-M | | 162 | 801 | 15063 | 8 | 70 | 4 | 5 | 6 | 116 | 6 | 1_0_2 | . | . | . | . | . | | |
| SPRLISCIII2465-24BR | ERS27214931 | 2024 | MAN | CAT | | 162 | 801 | 801 | 8 | 70 | 4 | 1 | 6 | 116 | 6 | 1_0_2 | . | . | . | . | . | | |
| SPRLISCIII2471-23 | ERS27214932 | 2023 | MAN | MUR | | 162 | 801 | 801 | 8 | 70 | 4 | 1 | 6 | 116 | 6 | 1_0_2 | . | . | . | . | . | | |
| SPRLISCIII2484-22 | ERS27214933 | 2022 | MAN | CAT | | 27 | 205 | 205 | 10 | 5 | 4 | 5 | 13 | 10 | 18 | 0_0_3 | . | . | . | . | . | | PI-1 |
| SPRLISCIII2507-09 | ERS27214934 | 2009 | MAN | GAL | | NA | 1866 | 1866 | 1 | 11 | 1 | 15 | 103 | 1 | 70 | 0_0_0 | . | . | . | . | . | | |
| SPRLISCIII2546-23 | ERS27214935 | 2023 | MAN | AND | SEV | 162 | 801 | 15063 | 8 | 70 | 4 | 5 | 6 | 116 | 6 | 1_0_2 | . | . | . | . | . | | |
| SPRLISCIII2548-19 | ERS27214936 | 2019 | MAN | CAT | | 182 | 1729 | 1729 | 1 | 5 | 1 | 1 | 15 | 169 | 14 | 2_4_2 | . | . | . | . | . | | PI-1 |
| SPRLISCIII2551-19 | ERS27214937 | 2019 | MAN | CAT | | 182 | 1729 | 1729 | 1 | 5 | 1 | 1 | 15 | 169 | 14 | 2_4_2 | . | . | . | . | . | | PI-1 |
| SPRLISCIII2556-22 | ERS27214938 | 2022 | WOMAN | AND | SEV | 162 | 801 | 15063 | 8 | 70 | 4 | 5 | 6 | 116 | 6 | 1_0_2 | . | . | . | . | . | | |
| SPRLISCIII2557-15 | ERS27214939 | 2015 | MAN | CAT | | 27 | 205 | 205 | 10 | 5 | 4 | 5 | 13 | 10 | 18 | 0_0_3 | . | . | . | . | . | | PI-1 |
| SPRLISCIII2557-22 | ERS27214940 | 2022 | MAN | AND | SEV | 162 | 801 | 15063 | 8 | 70 | 4 | 5 | 6 | 116 | 6 | 1_0_2 | . | . | . | . | . | | |
| SPRLISCIII2570-24 | ERS27214941 | 2024 | MAN | AND | | 162 | 801 | 15063 | 8 | 70 | 4 | 5 | 6 | 116 | 6 | 1_0_2 | . | . | . | . | . | | |
| SPRLISCIII2577-VNORMAL-24 | ERS27214942 | 2024 | WOMAN | AND | | 162 | 801 | 15063 | 8 | 70 | 4 | 5 | 6 | 116 | 6 | 1_0_2 | . | . | . | . | . | | |

| | | | | | | | | | | | | | | | | | | | | | | | |
|---|---|---|---|---|---|---|---|---|---|---|---|---|---|---|---|---|---|---|---|---|---|---|---|
| SPRLISCIII2585-23 | ERS27214943 | 2023 | MAN | AST | | 162 | 801 | 801 | 8 | 70 | 4 | 1 | 6 | 116 | 6 | 1_0_2 | . | . | . | . | . | | |
| SPRLISCIII2590-24 | ERS27214944 | 2024 | MAN | AND | | 70 | 259 | 7026 | 24 | 5 | 1 | 1 | 15 | 12 | 43 | 2_44_2 | . | . | . | . | . | | PI-1 |
| SPRLISCIII2594-19 | ERS27214945 | 2019 | MAN | CAN | | 162 | 801 | 801 | 8 | 70 | 4 | 1 | 6 | 116 | 6 | 1_0_2 | . | . | . | . | . | | |
| SPRLISCIII2596-19 | ERS27214946 | 2019 | WOMAN | VAL | | NA | 1866 | 1866 | 1 | 11 | 1 | 15 | 103 | 1 | 70 | 0_0_0 | . | . | . | . | . | | |
| SPRLISCIII2600-19 | ERS27214947 | 2019 | MAN | CAN | | 162 | 801 | 801 | 8 | 70 | 4 | 1 | 6 | 116 | 6 | 1_0_2 | . | . | . | . | . | | |
| SPRLISCIII2620-23 | ERS27214948 | 2023 | MAN | VAS | | 27 | 205 | 205 | 10 | 5 | 4 | 5 | 13 | 10 | 18 | 0_0_3 | . | . | . | . | . | | PI-1 |
| SPRLISCIII2622-23 | ERS27214949 | 2023 | WOMAN | EXT | | 162 | 801 | 15063 | 8 | 70 | 4 | 5 | 6 | 116 | 6 | 1_0_2 | . | . | . | . | . | | |
| SPRLISCIII2629-22 | ERS27214950 | 2022 | MAN | AND | SEV | 162 | 801 | 15063 | 8 | 70 | 4 | 5 | 6 | 116 | 6 | 1_0_2 | . | . | . | . | . | | |
| SPRLISCIII2630-15 | ERS27214951 | 2015 | MAN | VAS | | 162 | 801 | 1222 | 8 | 32 | 4 | 1 | 6 | 116 | 6 | 23_40_2 | . | . | . | . | . | | |
| SPRLISCIII2630-24B | ERS27214952 | 2024 | MAN | AND | SEV | 162 | 801 | 15063 | 8 | 70 | 4 | 5 | 6 | 116 | 6 | 1_0_2 | . | . | . | . | . | | |
| SPRLISCIII2632-19 | ERS27214953 | 2019 | MAN | CAN | | 162 | 801 | 801 | 8 | 70 | 4 | 1 | 6 | 116 | 6 | 1_0_2 | . | . | . | . | . | | |
| SPRLISCIII2636-24 | ERS27214954 | 2024 | MAN | AND | SEV | 162 | 801 | 15063 | 8 | 70 | 4 | 5 | 6 | 116 | 6 | 1_0_2 | . | . | . | . | . | | |
| SPRLISCIII2639-22 | ERS27214955 | 2022 | WOMAN | MUR | | 162 | 801 | 15063 | 8 | 70 | 4 | 5 | 6 | 116 | 6 | 1_0_2 | . | . | . | . | . | | |
| SPRLISCIII2643-22 | ERS27214956 | 2022 | MAN | AND | | 162 | 801 | 15063 | 8 | 70 | 4 | 5 | 6 | 116 | 6 | 1_0_2 | . | . | . | . | . | | |
| SPRLISCIII2654-18 | ERS27214957 | 2018 | WOMAN | CAT | | 331 | 1469 | 1469 | 8 | 88 | 103 | 18 | 46 | 142 | 161 | 12_2_418 | . | . | . | . | . | PPH015 | |
| SPRLISCIII2654-23 | ERS27214958 | 2023 | WOMAN | ARA | | 427 | 10172 | 10172 | 1 | 8 | 9 | 15 | 6 | 58 | 70 | 3_0_0 | . | . | . | . | . | | |
| SPRLISCIII2671-24 | ERS27214959 | 2024 | MAN | VAS | | 182 | 2941 | 9197 | 97 | 5 | 1 | 226 | 36 | 1 | 14 | 2_4_2 | . | . | . | . | . | | PI-1 |
| SPRLISCIII2683-23 | ERS27214960 | 2023 | MAN | AND | SEV | 162 | 801 | 15063 | 8 | 70 | 4 | 5 | 6 | 116 | 6 | 1_0_2 | . | . | . | . | . | | |
| SPRLISCIII2691-24 | ERS27214961 | 2024 | WOMAN | CAN | | NA | 1866 | 1866 | 1 | 11 | 1 | 15 | 103 | 1 | 70 | 0_0_0 | . | . | . | . | . | | |
| SPRLISCIII2693-24 | ERS27214962 | 2024 | MAN | CAT | | 162 | 801 | 13022 | 8 | 32 | 4 | 1 | 6 | 4 | 6 | NEW_0_2 | . | . | . | . | . | | |
| SPRLISCIII2701-24 | ERS27214963 | 2024 | MAN | CAT | | 162 | 801 | 13022 | 8 | 32 | 4 | 1 | 6 | 4 | 6 | NEW_0_2 | . | . | . | . | . | | |
| SPRLISCIII2707-23 | ERS27214964 | 2023 | MAN | AND | SEV | 162 | 801 | 15063 | 8 | 70 | 4 | 5 | 6 | 116 | 6 | 1_0_2 | . | . | . | . | . | | |

| | | | | | | | | | | | | | | | | | | | | | | | |
|---|---|---|---|---|---|---|---|---|---|---|---|---|---|---|---|---|---|---|---|---|---|---|---|
| SPRLISCIII2718-09 | ERS27214965 | 2009 | MAN | MAD | | 27 | 899 | 247 | 16 | 13 | 4 | 5 | 6 | 10 | 14 | 0_0_3 | . | . | . | . | . | | PI-1 |
| SPRLISCIII2722-24 | ERS27214966 | 2024 | MAN | CAT | | 162 | 801 | 13022 | 8 | 32 | 4 | 1 | 6 | 4 | 6 | NEW_0_2 | . | . | . | . | . | | |
| SPRLISCIII2749-24 | ERS27214967 | 2024 | MAN | MUR | | 27 | 899 | 247 | 16 | 13 | 4 | 5 | 6 | 10 | 14 | 0_0_3 | . | . | . | . | . | | PI-1 |
| SPRLISCIII2759-23 | ERS27214968 | 2023 | MAN | EXT | | 162 | 801 | 15063 | 8 | 70 | 4 | 5 | 6 | 116 | 6 | 1_0_2 | . | . | . | . | . | | |
| SPRLISCIII2762-15 | ERS27214969 | 2015 | MAN | C-M | | 162 | 801 | 1222 | 8 | 32 | 4 | 1 | 6 | 116 | 6 | NEW_0_2 | . | . | . | . | . | | |
| SPRLISCIII2778-24V | ERS27214970 | 2024 | MAN | NAV | | 162 | 801 | 15063 | 8 | 70 | 4 | 5 | 6 | 116 | 6 | 1_0_2 | . | . | . | . | . | | |
| SPRLISCIII2802-24 | ERS27214971 | 2024 | WOMAN | AND | | 162 | 801 | 15063 | 8 | 70 | 4 | 5 | 6 | 116 | 6 | 1_0_2 | . | . | . | . | . | | |
| SPRLISCIII2803-18 | ERS27214972 | 2018 | MAN | CAT | | 162 | 801 | 13022 | 8 | 32 | 4 | 1 | 6 | 4 | 6 | NEW_0_2 | . | . | . | . | . | | |
| SPRLISCIII2817-23 | ERS27214973 | 2023 | MAN | CAT | | 162 | 801 | 13022 | 8 | 32 | 4 | 1 | 6 | 4 | 6 | NEW_0_2 | . | . | . | . | . | | |
| SPRLISCIII2820-24 | ERS27214974 | 2024 | MAN | CAT | | 162 | 19282 | 19282 | 244 | 9 | 4 | 1 | 6 | 116 | 6 | NEW_0_2 | . | . | . | . | *tet*(M)_1 | Tn*916*-family | |
| SPRLISCIII2837-23 | ERS27214975 | 2023 | MAN | CAT | | 427 | 10172 | 10172 | 1 | 8 | 9 | 15 | 6 | 58 | 70 | 3_0_0 | . | . | . | . | . | | |
| SPRLISCIII2846-24 | ERS27214976 | 2024 | WOMAN | AND | | 162 | 801 | 15063 | 8 | 70 | 4 | 5 | 6 | 116 | 6 | 1_0_2 | . | . | . | . | . | | |
| SPRLISCIII2859-24V | ERS27214977 | 2024 | MAN | AND | | 162 | 801 | 15063 | 8 | 70 | 4 | 5 | 6 | 116 | 6 | 1_0_2 | . | . | . | . | . | | |
| SPRLISCIII2881-23 | ERS27214978 | 2023 | WOMAN | AND | SEV | 162 | 801 | 15063 | 8 | 70 | 4 | 5 | 6 | 116 | 6 | 1_0_2 | . | . | . | . | . | | |
| SPRLISCIII2881-24V | ERS27214979 | 2024 | MAN | BAL | | NA | 1866 | 1866 | 1 | 11 | 1 | 15 | 103 | 1 | 70 | 0_0_0 | . | . | . | . | . | | |
| SPRLISCIII2886-23 | ERS27214980 | 2023 | WOMAN | AND | | 162 | 801 | 15063 | 8 | 70 | 4 | 5 | 6 | 116 | 6 | 1_0_2 | . | . | . | . | . | | |
| SPRLISCIII2887-23 | ERS27214981 | 2023 | MAN | AND | | 162 | 801 | 15063 | 8 | 70 | 4 | 5 | 6 | 116 | 6 | 1_0_2 | . | . | . | . | . | | |
| SPRLISCIII2888-24 | ERS27214982 | 2024 | MAN | AND | | 182 | 1729 | 1729 | 1 | 5 | 1 | 1 | 15 | 169 | 14 | 2_4_2 | . | . | . | . | . | | PI-1 |
| SPRLISCIII2889-24B | ERS27214983 | 2024 | MAN | AND | SEV | 162 | 801 | 15063 | 8 | 70 | 4 | 5 | 6 | 116 | 6 | 1_0_2 | . | . | . | . | . | | |
| SPRLISCIII2892-24 | ERS27214984 | 2024 | MAN | AND | SEV | 162 | 801 | 15063 | 8 | 70 | 4 | 5 | 6 | 116 | 6 | 1_0_2 | . | . | . | . | . | | |
| SPRLISCIII2893-24 | ERS27214985 | 2024 | MAN | AND | | 162 | 801 | 15063 | 8 | 70 | 4 | 5 | 6 | 116 | 6 | 1_0_2 | . | . | . | . | . | | |
| SPRLISCIII2922-24 | ERS27214986 | 2024 | MAN | EXT | | 162 | 801 | 15063 | 8 | 70 | 4 | 5 | 6 | 116 | 6 | 1_0_2 | . | . | . | . | . | | |

| | | | | | | | | | | | | | | | | | | | | | | | | |
|---|---|---|---|---|---|---|---|---|---|---|---|---|---|---|---|---|---|---|---|---|---|---|---|---|
| SPRLISCIII2923-23 | ERS27214987 | 2023 | MAN | AND | SEV | 162 | 801 | 801 | 8 | 70 | 4 | 1 | 6 | 116 | 6 | 1_0_2 | . | . | *mef*(A)_2 | *msr*(D)_2 | . | ND | | |
| SPRLISCIII2925-23 | ERS27214988 | 2023 | MAN | AND | SEV | 162 | 801 | 15063 | 8 | 70 | 4 | 5 | 6 | 116 | 6 | 1_0_2 | . | . | . | . | . | | | |
| SPRLISCIII2926-23 | ERS27214989 | 2023 | MAN | AND | SEV | 162 | 801 | 15063 | 8 | 70 | 4 | 5 | 6 | 116 | 6 | 1_0_2 | . | . | . | . | . | | | |
| SPRLISCIII2928-23 | ERS27214990 | 2023 | WOMAN | AND | SEV | 162 | 801 | 801 | 8 | 70 | 4 | 1 | 6 | 116 | 6 | 1_0_2 | . | . | *mef*(A)_2 | *msr*(D)_2 | . | ND | | |
| SPRLISCIII2938-24 | ERS27214991 | 2024 | MAN | ARA | | 162 | 801 | 15063 | 8 | 70 | 4 | 5 | 6 | 116 | 6 | 1_0_2 | . | . | . | . | . | | | |
| SPRLISCIII2947-23 | ERS27214992 | 2023 | WOMAN | AND | SEV | 162 | 801 | 15063 | 8 | 70 | 4 | 5 | 6 | 116 | 6 | 1_0_2 | . | . | . | . | . | | | |
| SPRLISCIII2989-24V | ERS27214993 | 2024 | MAN | AND | SEV | 162 | 801 | 15063 | 8 | 70 | 4 | 5 | 6 | 116 | 6 | 1_0_2 | . | . | . | . | . | | | |
| SPRLISCIII2995-24 | ERS27214994 | 2024 | MAN | EXT | | 162 | 801 | 15063 | 8 | 70 | 4 | 5 | 6 | 116 | 6 | 1_0_2 | . | . | . | . | . | | | |
| SPRLISCIII3004-24 | ERS27214995 | 2024 | MAN | CAN | | 162 | 801 | 801 | 8 | 70 | 4 | 1 | 6 | 116 | 6 | 1_0_2 | . | . | . | . | . | | | |
| SPRLISCIII3016-23 | ERS27214996 | 2023 | MAN | CAT | | 162 | 801 | 13022 | 8 | 32 | 4 | 1 | 6 | 4 | 6 | NEW_0_2 | . | . | . | . | . | | | |
| SPRLISCIII3021-23 | ERS27214997 | 2023 | MAN | CAT | | 162 | 801 | 13022 | 8 | 32 | 4 | 1 | 6 | 4 | 6 | NEW_0_2 | . | . | . | . | . | | | |
| SPRLISCIII3042-19 | ERS27214998 | 2019 | WOMAN | GAL | | 162 | 801 | 801 | 8 | 70 | 4 | 1 | 6 | 116 | 6 | 1_0_2 | . | . | . | . | . | | | |
| SPRLISCIII3048-24 | ERS27214999 | 2024 | MAN | CAN | | 27 | 205 | 205 | 10 | 5 | 4 | 5 | 13 | 10 | 18 | 0_0_3 | . | . | . | . | . | | | PI-1 |
| SPRLISCIII3050-19 | ERS27215000 | 2019 | MAN | GAL | | 162 | 801 | 15063 | 8 | 70 | 4 | 5 | 6 | 116 | 6 | 1_0_2 | . | . | . | . | . | | | |
| SPRLISCIII3056-09 | ERS27215001 | 2009 | MAN | AND | | 27 | 899 | 247 | 16 | 13 | 4 | 5 | 6 | 10 | 14 | 0_0_3 | . | . | . | . | . | | | PI-1 |
| SPRLISCIII3056-23 | ERS27215002 | 2023 | MAN | MUR | | NA | 1866 | NEW | 1 | 11 | 1 | ~486 | 103 | 1 | 70 | 0_0_0 | . | . | . | . | . | | | |
| SPRLISCIII3070-23 | ERS27215003 | 2023 | WOMAN | AND | | 162 | 801 | 15063 | 8 | 70 | 4 | 5 | 6 | 116 | 6 | 1_0_2 | . | . | . | . | . | | | |
| SPRLISCIII3104-15 | ERS27215004 | 2015 | MAN | MUR | | 27 | 244 | 244 | 16 | 2 | 4 | 1 | 6 | 10 | 18 | 0_0_3 | *cat*(pC194)_1 | *erm*(B)_18 | . | . | tet(M)_8 | Tn*916*-family | | PI-1 |
| SPRLISCIII3110-24 | ERS27215005 | 2024 | MAN | AND | | 162 | 801 | 15063 | 8 | 70 | 4 | 5 | 6 | 116 | 6 | 1_0_2 | . | . | . | . | . | | | |
| SPRLISCIII3142-24 | ERS27215006 | 2024 | MAN | C-M | | 27 | 205 | 205 | 10 | 5 | 4 | 5 | 13 | 10 | 18 | 0_0_3 | . | . | . | . | . | | PPH010 | PI-1 |
| SPRLISCIII3146-15 | ERS27215007 | 2015 | MAN | CAT | | 162 | 801 | 801 | 8 | 70 | 4 | 1 | 6 | 116 | 6 | 1_0_2 | . | . | . | . | . | | | |
| SPRLISCIII3147-24V | ERS27215008 | 2024 | WOMAN | CAT | | 27 | 899 | 247 | 16 | 13 | 4 | 5 | 6 | 10 | 14 | 0_0_3 | . | . | . | . | . | | | PI-1 |

| | | | | | | | | | | | | | | | | | | | | | | | |
|---|---|---|---|---|---|---|---|---|---|---|---|---|---|---|---|---|---|---|---|---|---|---|---|
| SPRLISCIII3152-23 | ERS27215009 | 2023 | MAN | AST | | 162 | 801 | 801 | 8 | 70 | 4 | 1 | 6 | 116 | 6 | 1_0_2 | . | . | . | . | . | | |
| SPRLISCIII3162-19 | ERS27215010 | 2019 | WOMAN | VAS | | 162 | 801 | 1222 | 8 | 32 | 4 | 1 | 6 | 116 | 6 | 23_40_2 | . | . | . | . | . | | |
| SPRLISCIII3162-24 | ERS27215011 | 2024 | MAN | AND | SEV | 162 | 801 | 15063 | 8 | 70 | 4 | 5 | 6 | 116 | 6 | 1_0_2 | . | . | . | . | . | | |
| SPRLISCIII3168-23 | ERS27215012 | 2023 | MAN | AND | | 162 | 801 | 15063 | 8 | 70 | 4 | 5 | 6 | 116 | 6 | 1_0_2 | . | . | . | . | . | | |
| SPRLISCIII3174-24 | ERS27215013 | 2024 | MAN | AND | SEV | 162 | 801 | 15063 | 8 | 70 | 4 | 5 | 6 | 116 | 6 | 1_0_2 | . | . | . | . | . | | |
| SPRLISCIII3178-24 | ERS27215014 | 2024 | MAN | AND | SEV | 162 | 801 | 15063 | 8 | 70 | 4 | 5 | 6 | 116 | 6 | 1_0_2 | . | . | . | . | . | | |
| SPRLISCIII3183-15 | ERS27215015 | 2015 | MAN | ARA | | 27 | 205 | 205 | 10 | 5 | 4 | 5 | 13 | 10 | 18 | 0_0_3 | . | . | . | . | . | | PI-1 |
| SPRLISCIII3184-24 | ERS27215016 | 2024 | MAN | AND | SEV | 162 | 801 | 15063 | 8 | 70 | 4 | 5 | 6 | 116 | 6 | 1_0_2 | . | . | . | . | . | | |
| SPRLISCIII3192-09 | ERS27215017 | 2009 | MAN | EXT | | 27 | 244 | 244 | 16 | 2 | 4 | 1 | 6 | 10 | 18 | 0_0_3 | . | . | . | . | . | | PI-1 |
| SPRLISCIII3199-24V | ERS27215018 | 2024 | MAN | CAN | | 162 | 801 | 15063 | 8 | 70 | 4 | 5 | 6 | 116 | 6 | 1_0_2 | . | . | . | . | . | | |
| SPRLISCIII3236-24 | ERS27215019 | 2024 | WOMAN | MUR | | 27 | 899 | 247 | 16 | 13 | 4 | 5 | 6 | 10 | 14 | 0_0_3 | . | . | . | . | . | | PI-1 |
| SPRLISCIII3241-23 | ERS27215020 | 2023 | MAN | EXT | | 162 | 801 | 15063 | 8 | 70 | 4 | 5 | 6 | 116 | 6 | 1_0_2 | . | . | . | . | . | | |
| SPRLISCIII3244-24 | ERS27215021 | 2024 | MAN | MAD | | 162 | 801 | 15063 | 8 | 70 | 4 | 5 | 6 | 116 | 6 | 1_0_2 | . | . | . | . | . | | |
| SPRLISCIII3250-15 | ERS27215022 | 2015 | WOMAN | CAT | | 27 | 205 | 205 | 10 | 5 | 4 | 5 | 13 | 10 | 18 | 0_0_3 | . | . | . | . | . | | PI-1 |
| SPRLISCIII3253-24 | ERS27215023 | 2024 | MAN | AND | | 27 | 899 | 247 | 16 | 13 | 4 | 5 | 6 | 10 | 14 | 0_0_3 | . | . | . | . | . | | PI-1 |
| SPRLISCIII3254-24V | ERS27215024 | 2024 | WOMAN | AND | | 162 | 801 | 15063 | 8 | 70 | 4 | 5 | 6 | 116 | 6 | 1_0_2 | . | . | . | . | . | | |
| SPRLISCIII3260-19 | ERS27215025 | 2019 | MAN | MUR | | 162 | 801 | 801 | 8 | 70 | 4 | 1 | 6 | 116 | 6 | 1_0_2 | . | . | *mef*(A)_2 | *msr*(D)_2 | . | ND | |
| SPRLISCIII3266-24 | ERS27215026 | 2024 | MAN | AND | SEV | 162 | 801 | 15063 | 8 | 70 | 4 | 5 | 6 | 116 | 6 | 1_0_2 | . | . | . | . | . | | |
| SPRLISCIII3273-15 | ERS27215027 | 2015 | WOMAN | AND | | 27 | 205 | 205 | 10 | 5 | 4 | 5 | 13 | 10 | 18 | 0_0_3 | . | . | . | . | . | | PI-1 |
| SPRLISCIII3293-23 | ERS27215028 | 2023 | WOMAN | AND | SEV | 162 | 801 | 15063 | 8 | 70 | 4 | 5 | 6 | 116 | 6 | 1_0_2 | . | . | . | . | . | | |
| SPRLISCIII3295-23 | ERS27215029 | 2023 | MAN | AND | SEV | 162 | 801 | 15063 | 8 | 70 | 4 | 5 | 6 | 116 | 6 | 1_0_2 | . | . | . | . | . | | |
| SPRLISCIII3296-23 | ERS27215030 | 2023 | MAN | AND | SEV | 162 | 801 | 15063 | 8 | 70 | 4 | 5 | 6 | 116 | 6 | 1_0_2 | . | . | . | . | . | | |

| | | | | | | | | | | | | | | | | | | | | | | |
|---|---|---|---|---|---|---|---|---|---|---|---|---|---|---|---|---|---|---|---|---|---|---|
| SPRLISCIII3323-09 | ERS27215031 | 2009 | MAN | GAL | | 27 | 899 | 246 | 16 | 13 | 4 | 5 | 6 | 10 | 18 | 0_0_3 | . | . | . | . | . | | PI-1 |
| SPRLISCIII3333-23 | ERS27215032 | 2023 | WOMAN | C-M | | 162 | 801 | 15063 | 8 | 70 | 4 | 5 | 6 | 116 | 6 | 1_0_2 | . | . | . | . | . | | |
| SPRLISCIII3376-23 | ERS27215033 | 2023 | WOMAN | AND | SEV | 162 | 801 | 15063 | 8 | 70 | 4 | 5 | 6 | 116 | 6 | 1_0_2 | . | . | . | . | . | | |
| SPRLISCIII3384-09 | ERS27215034 | 2009 | MAN | CAT | | 27 | 4829 | 4829 | 16 | 17 | 4 | 1 | 135 | 10 | 14 | 0_0_0 | . | . | . | . | *tet*(M)_1 | Tn*916*-family | PI-1 |
| SPRLISCIII3386-19 | ERS27215035 | 2019 | WOMAN | AND | | 70 | 259 | 7026 | 24 | 5 | 1 | 1 | 15 | 12 | 43 | 2_44_2 | . | . | . | . | . | | PI-1 |
| SPRLISCIII3388-23 | ERS27215036 | 2023 | WOMAN | AND | SEV | 162 | 801 | 15063 | 8 | 70 | 4 | 5 | 6 | 116 | 6 | 1_0_2 | . | . | . | . | . | | |
| SPRLISCIII3408-23 | ERS27215037 | 2023 | MAN | AND | | 162 | 801 | 15063 | 8 | 70 | 4 | 5 | 6 | 116 | 6 | 1_0_2 | . | . | . | . | . | | |
| SPRLISCIII3421-23 | ERS27215038 | 2023 | MAN | MAD | | 162 | 801 | 801 | 8 | 70 | 4 | 1 | 6 | 116 | 6 | 1_0_2 | . | . | *mef*(A)_2 | *msr*(D)_2 | . | ND | |
| SPRLISCIII3424-23 | ERS27215039 | 2023 | WOMAN | MAD | | 27 | 899 | 247 | 16 | 13 | 4 | 5 | 6 | 10 | 14 | 0_0_3 | . | . | . | . | . | | PI-1 |
| SPRLISCIII3446-23 | ERS27215040 | 2023 | MAN | AND | SEV | 162 | 801 | 15063 | 8 | 70 | 4 | 5 | 6 | 116 | 6 | 1_0_2 | . | . | . | . | . | | |
| SPRLISCIII3461-09 | ERS27215041 | 2009 | WOMAN | CAT | | 27 | 205 | 205 | 10 | 5 | 4 | 5 | 13 | 10 | 18 | 0_0_3 | . | . | . | . | . | | PI-1 |
| SPRLISCIII3466-23 | ERS27215042 | 2023 | WOMAN | AND | SEV | 162 | 801 | 15063 | 8 | 70 | 4 | 5 | 6 | 116 | 6 | 1_0_2 | . | . | . | . | . | | |
| SPRLISCIII3469-23 | ERS27215043 | 2023 | MAN | AND | SEV | 162 | 801 | 15063 | 8 | 70 | 4 | 5 | 6 | 116 | 6 | 1_0_2 | . | . | . | . | . | | |
| SPRLISCIII3470-23 | ERS27215044 | 2023 | WOMAN | AND | SEV | 162 | 801 | 15063 | 8 | 70 | 4 | 5 | 6 | 116 | 6 | 1_0_2 | . | . | . | . | . | | |
| SPRLISCIII3471-23 | ERS27215045 | 2023 | MAN | AND | SEV | 162 | 801 | 15063 | 8 | 70 | 4 | 5 | 6 | 116 | 6 | 1_0_2 | . | . | . | . | . | | |
| SPRLISCIII3472-23 | ERS27215046 | 2023 | MAN | CAT | | 162 | 801 | 1222 | 8 | 32 | 4 | 1 | 6 | 116 | 6 | NEW_0_2 | . | . | . | . | . | | |
| SPRLISCIII3474-23 | ERS27215047 | 2023 | MAN | AND | SEV | 162 | 801 | 15063 | 8 | 70 | 4 | 5 | 6 | 116 | 6 | 1_0_2 | . | . | . | . | . | | |
| SPRLISCIII3507-23 | ERS27215048 | 2023 | MAN | AND | | 162 | 801 | 15063 | 8 | 70 | 4 | 5 | 6 | 116 | 6 | 1_0_2 | . | . | . | . | . | | |
| SPRLISCIII3516-23 | ERS27215049 | 2023 | MAN | EXT | | 162 | 801 | 15063 | 8 | 70 | 4 | 5 | 6 | 116 | 6 | 1_0_2 | . | . | . | . | . | | |
| SPRLISCIII3529-23 | ERS27215050 | 2023 | MAN | MAD | | 162 | 801 | 15063 | 8 | 70 | 4 | 5 | 6 | 116 | 6 | 1_0_2 | . | . | . | . | . | | |
| SPRLISCIII3556-23 | ERS27215051 | 2023 | MAN | AND | SEV | 162 | 801 | 15063 | 8 | 70 | 4 | 5 | 6 | 116 | 6 | 1_0_2 | . | . | . | . | . | | |
| SPRLISCIII3563-23 | ERS27215052 | 2023 | MAN | AND | SEV | 162 | 801 | 15063 | 8 | 70 | 4 | 5 | 6 | 116 | 6 | 1_0_2 | . | . | . | . | . | | |

| | | | | | | | | | | | | | | | | | | | | | | | |
|---|---|---|---|---|---|---|---|---|---|---|---|---|---|---|---|---|---|---|---|---|---|---|---|
| SPRLISCIII3571-09 | ERS27215053 | 2009 | MAN | NAV | | NA | 1866 | 1866 | 1 | 11 | 1 | 15 | 103 | 1 | 70 | 0_0_0 | . | . | . | . | . | | |
| SPRLISCIII3602-09 | ERS27215054 | 2009 | MAN | C-L | | 8 | 289 | 1637 | 10 | 13 | 53 | 5 | 13 | 10 | 18 | 0_0_3 | . | . | . | . | . | | PI-1 |
| SPRLISCIII3610-09 | ERS27215055 | 2009 | MAN | GAL | | 70 | 1221 | 4795 | 7 | 5 | 1 | 5 | 15 | 12 | 8 | 2_0_3 | . | . | . | . | . | | PI-1 |
| SPRLISCIII3620-23 | ERS27215056 | 2023 | MAN | AND | | 162 | 801 | 15063 | 8 | 70 | 4 | 5 | 6 | 116 | 6 | 1_0_2 | . | . | . | . | . | | |
| SPRLISCIII3630-23 | ERS27215057 | 2023 | MAN | AND | SEV | 162 | 801 | 15063 | 8 | 70 | 4 | 5 | 6 | 116 | 6 | 1_0_2 | . | . | . | . | . | | |
| SPRLISCIII3634-23 | ERS27215058 | 2023 | MAN | AND | SEV | 162 | 801 | 15063 | 8 | 70 | 4 | 5 | 6 | 116 | 6 | 1_0_2 | . | . | . | . | . | | |
| SPRLISCIII3650-23 | ERS27215059 | 2023 | MAN | C-M | | 27 | 899 | 247 | 16 | 13 | 4 | 5 | 6 | 10 | 14 | 0_0_3 | . | . | . | . | . | | PI-1 |
| SPRLISCIII3653-23 | ERS27215060 | 2023 | MAN | MAD | | 27 | 899 | 247 | 16 | 13 | 4 | 5 | 6 | 10 | 14 | 0_0_3 | . | . | . | . | . | | PI-1 |
| SPRLISCIII3702-09 | ERS27215061 | 2009 | WOMAN | GAL | | 27 | 899 | 247 | 16 | 13 | 4 | 5 | 6 | 10 | 14 | 0_0_3 | . | . | . | . | . | | PI-1 |
| SPRLISCIII3711-09 | ERS27215062 | 2009 | MAN | AND | | 70 | 1221 | 4795 | 7 | 5 | 1 | 5 | 15 | 12 | 8 | 2_0_3 | . | . | . | . | . | | PI-1 |
| SPRLISCIII3792-09 | ERS27215063 | 2009 | MAN | AND | | 27 | 899 | 247 | 16 | 13 | 4 | 5 | 6 | 10 | 14 | 0_0_3 | . | . | . | . | *tet*(M)_12 | Tn*916*-family | PI-1 |
| SPRLISCIII3844-09 | ERS27215064 | 2009 | MAN | CAT | | 27 | 205 | 205 | 10 | 5 | 4 | 5 | 13 | 10 | 18 | 0_0_3 | . | . | . | . | . | | PI-1 |
| SPRLISCIII3982-09 | ERS27215065 | 2009 | MAN | CAT | | 27 | 205 | 205 | 10 | 5 | 4 | 5 | 13 | 10 | 18 | 0_0_3 | . | . | . | . | . | | PI-1 |
| SPRLISCIII3992-09 | ERS27215066 | 2009 | WOMAN | AST | | NA | 1866 | 1866 | 1 | 11 | 1 | 15 | 103 | 1 | 70 | 0_0_0 | . | . | . | . | . | | |
| SPRLISCIII4115-09 | ERS27215067 | 2009 | MAN | CAT | | 70 | 1221 | 4795 | 7 | 5 | 1 | 5 | 15 | 12 | 8 | 2_0_3 | . | . | . | . | . | | PI-1 |
| SPRLISCIII4174-09 | ERS27215068 | 2009 | WOMAN | CAT | | 70 | 1221 | 4795 | 7 | 5 | 1 | 5 | 15 | 12 | 8 | 2_0_3 | . | . | . | . | . | | PI-1 |
| SPRLISCIII4245-09 | ERS27215069 | 2009 | MAN | CAT | | 70 | 1221 | 1221 | 7 | 5 | 1 | 5 | 15 | 12 | 14 | 2_4_3 | . | . | . | . | . | | PI-1 |
| SPRLISCIII4306-09 | ERS27215070 | 2009 | MAN | GAL | | 162 | 801 | 801 | 8 | 70 | 4 | 1 | 6 | 116 | 6 | 1_0_2 | . | . | . | . | . | | |
| SPRLISCIII4404-09 | ERS27215071 | 2009 | WOMAN | CAT | | 27 | 205 | 2333 | 10 | 5 | 4 | 5 | 13 | 194 | 18 | 0_0_3 | . | *erm*(B)_18 | . | . | *tet*(M)_12 | Tn*5252*-family | PI-1 |
| SPRLISCIII4442-09 | ERS27215072 | 2009 | WOMAN | AND | | 736 | 6864 | 6864 | 118 | 60 | 4 | 15 | 25 | 14 | 9 | 150_350_77 | . | . | . | . | . | | |
| SPRLISCIII4498-09 | ERS27215073 | 2009 | WOMAN | C-M | | 27 | 899 | 247 | 16 | 13 | 4 | 5 | 6 | 10 | 14 | 0_0_3 | . | . | . | . | . | | PI-1 |
| SPRLISCIII4542-09 | ERS27215074 | 2009 | MAN | NAV | | 27 | 899 | 246 | 16 | 13 | 4 | 5 | 6 | 10 | 18 | 0_0_3 | . | . | . | . | . | | PI-1 |

| | | | | | | | | | | | | | | | | | | | | | | |
|---|---|---|---|---|---|---|---|---|---|---|---|---|---|---|---|---|---|---|---|---|---|---|
| SPRLISCIII4549-09 | ERS27215075 | 2009 | MAN | CAT | 70 | 1221 | 4795 | 7 | 5 | 1 | 5 | 15 | 12 | 8 | 2_0_3 | . | . | . | . | . | | PI-1 |
| SPRLISCIII4657-09 | ERS27215076 | 2009 | MAN | NAV | NA | 1866 | 1866 | 1 | 11 | 1 | 15 | 103 | 1 | 70 | 0_0_0 | . | . | . | . | . | | |
| SPRLISCIII4665-09 | ERS27215077 | 2009 | WOMAN | AND | 27 | 205 | 205 | 10 | 5 | 4 | 5 | 13 | 10 | 18 | 0_0_3 | . | . | . | . | . | | PI-1 |
| SPRLISCIII4667-09 | ERS27215078 | 2009 | WOMAN | VAS | NA | 1866 | 1866 | 1 | 11 | 1 | 15 | 103 | 1 | 70 | 0_0_0 | . | . | . | . | . | | |
| SPRLISCIII4678-09 | ERS27215079 | 2009 | WOMAN | CAT | 27 | 899 | 246 | 16 | 13 | 4 | 5 | 6 | 10 | 18 | 0_0_3 | . | . | . | . | . | | PI-1 |
| SPRLISCIII4719-09 | ERS27215080 | 2009 | MAN | C-M | 27 | 205 | 206 | 10 | 5 | 17 | 5 | 13 | 10 | 18 | 0_0_3 | . | . | . | . | . | | PI-1 |
| SPRLISCIII4764-09 | ERS27215081 | 2009 | WOMAN | AND | 27 | 205 | 205 | 10 | 5 | 4 | 5 | 13 | 10 | 18 | 0_0_3 | . | . | . | . | . | | PI-1 |
| SPRLISCIII4807-09 | ERS27215082 | 2009 | MAN | NAV | 27 | 899 | 246 | 16 | 13 | 4 | 5 | 6 | 10 | 18 | 0_0_3 | . | . | . | . | . | | PI-1 |

| **Isolate** | **Genotype/Subtype** | **Assay** | **Reference** |
|---|---|---|---|
| SPRLICIII4-18 | *ST15063/CC801/GPSC162* | Adhesion (Detroit, A549 & CSE) | SPRL |
| SPRLICIII1754-18 | *ST15063/CC801/GPSC162* | Adhesion (Detroit, A549 & CSE) | SPRL |
| SPRLICIII1324-19 | *ST15063/CC801/GPSC162* | Adhesion (Detroit, A549 & CSE) | SPRL |
| SPRLICIII582-23 | *ST15063/CC801/GPSC162* | Adhesion (Detroit, A549, CSE & int. IV) | SPRL |
| SPRLICIII1179-23 | *ST15063/CC801/GPSC162* | Adhesion (Detroit, A549 & CSE) | SPRL |
| SPRLICIII1728-23 | *ST15063/CC801/GPSC162* | Adhesion (Detroit, A549 & CSE) | SPRL |
| SPRLICIII1972-23 | *ST15063/CC801/GPSC162* | Adhesion (Detroit, A549 & CSE) | SPRL |
| SPRLICIII724-18 | *ST205/CC205/GPSC27* | Adhesion (Detroit, A549, CSE & int. IV) | SPRL |
| SPRLICIII800-23 | *ST205/CC205/GPSC27* | Adhesion (Detroit, A549 & CSE) | SPRL |
| A/New Caledonia/20/1999 | *H1N1* | Int. With IV | Mount Sinai |
| A/Darwin/9/2021 | *H3N2* | Int. With IV | Mount Sinai |

**Supplemental Table S2. Strains used in different studies.** Clinical isolates of *Streptococcus pneumoniae* were selected from the WGS analysis. Adhesion refers to assays of pulmonary (A549) and nasopharyngeal (Detroit 562) epithelial cells adhesion, also with the presence of cigarette smoke extract (CSE) and influenza virus (int. IV).
